# Systematic review of the entomological impact of insecticide-treated nets evaluated using experimental hut trials in Africa

**DOI:** 10.1101/2021.04.07.21254306

**Authors:** Rebecca K Nash, Ben Lambert, Raphael N’Guessan, Corine Ngufor, Mark Rowland, Richard Oxborough, Sarah Moore, Patrick Tungu, Ellie Sherrard-Smith, Thomas S Churcher

## Abstract

**Background:** Resistance of anopheline mosquitoes to pyrethroid insecticides is spreading rapidly across sub-Saharan Africa, diminishing the efficacy of insecticide-treated nets (ITNs) — the primary tool for preventing malaria. The entomological efficacy of indoor vector control interventions can be measured in experimental hut trials (EHTs), which are specially designed to quantify the protection provided under controlled conditions. Experimental hut structures resemble local housing but allow collection of surviving exiting mosquitoes as well as dead or dying mosquitoes. There is a need to understand how the spread of resistance changes ITN efficacy and to elucidate factors influencing EHT results, including differences in experimental hut construction and design features, to support the development of novel vector control tools.

**Methods:** A comprehensive database of EHTs was compiled and summarised following a systematic review to identify all known trials investigating ITNs or indoor residual spraying (IRS) across sub-Saharan Africa. This analysis focuses on EHTs investigating ITNs and uses Bayesian statistical models to characterise the complex interaction between ITNs and mosquitoes, the variability between studies, and the impact of pyrethroid resistance.

**Results:** As resistance rises, the entomological efficacy of ITNs declines. They induce less mortality and are less likely to deter mosquitoes from entering huts. Despite this, ITNs continue to offer considerable personal protection by reducing mosquito feeding until resistance reaches high levels. There are clear associations between the different entomological impacts of ITNs, though there is still substantial variability between studies, some of which can be accounted for by hut design. The relationship between EHT outcomes and the level of resistance (as measured by discriminating dose bioassays) is highly uncertain.

**Conclusions:** The meta-analyses show that EHTs are an important reproducible assay for capturing the complex entomological efficacy of ITNs on blood-feeding mosquitoes. The impact of pyrethroid resistance on these measures appears broadly consistent across a wide geographical area once hut design is accounted for, suggesting results can be extrapolated beyond the sites where the trials were conducted. Further work is needed to understand factors influencing EHT outcomes and how the relationship between outcomes and resistance varies when different methods are used to assess the level of resistance in wild mosquito populations. This will allow more precise estimates of the efficacy of these important vector control tools.

## Background

Insecticide-treated bed nets (ITNs) are the most widely used entomological interventions against malaria. ITNs include conventionally-treated nets (CTNs), where nets are ‘dipped’ in insecticide solution, and more recent and widely used long-lasting insecticide treated nets (LLINs), where the insecticide is incorporated or coated into the net’s fibres and maintain effective levels for up to 3 years or more^1^. Between 2000 and 2015, over 450 million cases of malaria are estimated to have been averted by ITNs.^2^ Over 230 million nets were distributed across Africa in 2020 alone^3^. Indoor residual spraying (IRS) of insecticides on the walls of houses is also highly effective, although uptake and application is less widespread^2^.

The effectiveness of global malaria control is threatened by widespread resistance of mosquitoes to pyrethroids — the only insecticide class used on the vast majority of LLINs.^4–8^ Resistance can be quantified using discriminating-dose bioassays, which measure the survival of wild mosquitoes after exposure to insecticide.^9^ As the frequency and intensity of pyrethroid resistance spreads across sub-Saharan Africa^10^, new types of nets are being developed to meet the challenge. The World Health Organisation (WHO) has identified four putative classes of ITNs – pyrethroid only, pyrethroid plus the synergist piperonyl butoxide (PBO), pyrethroid plus non-pyrethroid insecticide (with a different mode of action), and pyrethroid plus insect growth regulator. Only two of these – pyrethroid and pyrethroid-PBO – are currently recommended by WHO. Several brands of nets have been developed within each of these ITN classes, some of which are still under evaluation to fully demonstrate their public health value.

The WHO must evaluate and endorse vector control products that fall into the recommended intervention class before they can be purchased by United Nations agencies^11,12^. This prequalification process assesses whether the intervention satisfies international standards for quality, safety and efficacy^12^. As part of the assessment, the entomological efficacy of products is evaluated in experimental hut trials (EHTs). EHTs are an important ‘real-life’ bioassay used in the development and evaluation of new vector control tools and new products within an existing WHO-recognised class that target the mosquito inside houses. They involve human volunteers sleeping in huts designed to replicate housing in the study area. Huts are specially constructed with baffles and traps to retain mosquitoes that enter, which allows the mosquitoes entering, exiting, blood-feeding, and dying, to be collected without human interference and counted the next morning. Huts can either be a control hut, typically allocated either an untreated net or no net at all, or an intervention hut with an indoor intervention, such as an ITN or IRS. To reduce bias due to hut locations within the site, interventions tend to be rotated if possible. The key indicators used to assess the efficacy of an ITN or IRS are intervention-induced mortality and blood-feeding inhibition, although many other factors that may have an epidemiological impact can also be investigated, such as the relative number caught in intervention huts compared to the control hut^13^.

EHTs allow different indoor vector control tools to be directly compared at the same time within the same trial. These studies are known to have substantial measurement error due to the complex interaction between wild free-flying mosquitoes, the interventions themselves, the volunteer behaviour or odour, and the environment^14^. The accuracy and generalisability of EHTs conducted at different times and places is less clear and is exacerbated by variability in factors such as the mosquito species in the study area and deliberate differences in hut construction, size or design. There are three main hut designs used in sub-Saharan Africa, including: West African, East African and Ifakara style huts^15–17^. Each design differs in features such as the size of hut, the number and shape of entrances for mosquitoes and the type of exit trap, all of which are likely to affect trial outcomes. Massue et al. ^18^ previously tried to quantify the effect of hut design by carrying out a trial at the same site using huts of each type concurrently, but due to low mosquito numbers, were unable to determine differences in intervention effectiveness between huts. More recently, Oumbouke et al.^19^ evaluated West African and Ifakara huts and found differences in behaviour of nuisance Culex mosquitoes, though the impact on anopheline mosquitoes is undetermined. All other EHT sites tend to contain huts of a single type making comparisons between hut designs difficult. This is exacerbated as EHT outcomes may also depend on other factors specific to the site which cannot be controlled, for example, the local species of mosquito. These confounding factors could also make disentangling the processes influencing EHT outcomes difficult.

EHTs provide an important tool for assessing the entomological impact of ITNs on pyrethroid resistant mosquitoes in controlled field conditions. Despite their key importance in evaluating some of the world’s most effective public health tools, the variability of EHTs have rarely been studied. Products tend to be evaluated in WHO trial sites in both east and west Africa to partly address this deficiency when evaluating a new product. The lack of replicate EHTs in the same region or differences in vector species is a further problem. Meta-analyses of EHT data testing specific products are increasingly used to compare interventions and parameterise mathematical models of the transmission dynamics of malaria^20–24^. This study conducts a systematic review of published literature, WHO documentation and other grey literature, to summarise all known EHTs of ITN or IRS interventions conducted in Africa. A systematic review of the entomological impact of IRS in EHTs was recently completed by Sherrard-Smith et al^21^, so this study focuses on ITNs and extends an earlier meta-analysis of EHTs comparing treated and untreated nets, which found no clear relationship between resistance and ITN efficacy^25^. Here, we use statistical models to characterise how increasing pyrethroid resistance in the mosquito population (as measured using discriminating dose bioassays) changes the entomological efficacy of ITNs and quantify the impact of hut design on trial outcomes.

## Methods

### Search Strategy

A database of all relevant EHTs was compiled following a systematic search based on PRISMA guidelines (see Supplementary Figure 1). The protocol for the review was prospectively registered on PROSPERO (CRD42019117858). Electronic databases MEDLINE, EMBASE, Global Health, Scopus, Web of Science and Google Scholar were searched using the strategy outlined in Supplementary Table 1. Searches were adapted for each database and followed the same structure where possible. The identified studies were then screened using the inclusion and exclusion criteria displayed in Supplementary Table 2. If the criteria were fulfilled, data were then extracted into piloted forms. In addition, data from unpublished studies were taken from WHO Pesticide Evaluation Scheme (WHOPES) working group meeting reports, and contact was made with authors who regularly conduct EHTs to acquire any further grey literature.

### Location of EHTs

The EHT sites were mapped using the ggspatial package in R^26^. If coordinates for the trial site were not explicitly stated in the study, then the relevant coordinates were obtained by either matching the sites to those where coordinates had been provided elsewhere, or by using the description of the study site to find coordinates in Google Maps™ or OpenStreetMap™. Trials using the Ifakara hut design (n=6) were removed from the ensuing analyses as the design was only identified at one site.

### Summary statistics

Analyses are restricted to EHTs evaluating ITNs, including conventionally treated nets (CTNs) and long-lasting insecticide treated nets (LLINs), as the entomological impact of IRS has recently been evaluated elsewhere^18^. ITNs are either pyrethroid-only (including deltamethrin, permethrin and alphacypermethrin insecticides), pyrethroid-PBO (pyrethroid combined with the synergist piperonyl butoxide (PBO)), non-pyrethroid (including insecticides such as chlorfenapyr and pyriproxyfen) or pyrethroid-combination (pyrethroid combined with an insecticide from an alternative insecticide class, such as the pyrrole, chlorfenapyr). EHTs generally report either the number or proportion of mosquitoes that fed, died, exited and the total number of mosquitoes collected, in each control or intervention arm of the trial. Mortality data is reported as the number of mosquitoes collected from a hut that die over the subsequent 24-hours. If the ITN incorporated the insecticide chlorfenapyr, then the mortality reported is usually 72-hour delayed mortality to be consistent with its mode of action. It is assumed that if a mosquito enters a hut with an intervention attempting to blood-feed, they can either exit without blood-feeding, successfully blood-feed (which we define as being blood-fed and alive) or die.

As in previous work, we aim to determine the association between (i) the level of resistance measured in the field and EHT mosquito survival (A1) and (ii) Mosquito survival and other outcomes measured in EHTs (A2).

### Pyrethroid resistance and ITN survival (A1)

We attempt to characterise the association between the level of pyrethroid resistance in the local mosquito population and mosquito survival in EHTs of pyrethroid-only ITNs. The fraction of mosquitoes surviving discriminating dosages of insecticide for 24-hours following exposure can act as an indicator of the level of pyrethroid resistance in the population^27,28^. Discriminating dose bioassays (hereby referred to as bioassays) include the WHO susceptibility test and the Centers for Disease Control bottle assay^29,30^. The level of mortality determined in these bioassays is known to vary depending on environmental factors^31^ (which were not systematically measured across EHTs), but they remain the most widely used method for assessing resistance in wild mosquitoes and were selected for analysis. All EHTs identified in the systematic review that evaluated pyrethroid-only ITNs were screened to identify those where concurrent bioassays were conducted on the same mosquito population. The analysis was then restricted to EHT mortality data for unwashed nets, and, if studies did not report the total numbers of mosquitoes tested in the bioassay, they were allocated a total of 50 mosquitoes (corresponding to half the standard number of mosquitoes tested) to reduce their weighting on the fit. If a trial took place before the year 2000, when mass ITN distribution campaigns began^2^ (n=2), there was assumed to be no pyrethroid resistance. They were allocated a total number of 50 mosquitoes tested in the bioassay and all were assumed to survive. Raw data on the number of mosquitoes tested and surviving were used, so they are not corrected for control mortality.

We modelled the number of mosquitoes that survived in the bioassay for trial 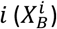 using a binomial model:

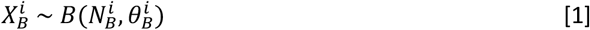

where 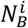 is the number of mosquitoes tested and 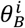 represents the probability of survival in the bioassay, which, in turn, is modelled:

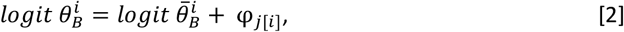

where *logit θ* = log[*θ*/(1 − *θ*)] and φ_*j*[*i*]_ is a location-level effect corresponding to the location, *j*, where trial *i* took place (i.e. there are potentially multiple trials per location). Here, 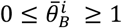 represents the estimate of bioassay survival when that trial was conducted after accounting for location-level variability. We set priors: 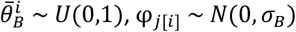 and *σ*_*B*_~*half N*(0,1).

The number of mosquitoes surviving in hut trials 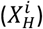 is characterised in a similar fashion:

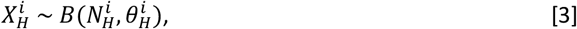

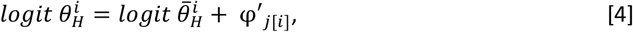

where 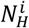 is the number of mosquitoes caught in the experimental hut for each trial arm *i*. As before, we specify the following priors on the location-specific effects: φ′_*j*[*i*]_ ~ *N*(0, *σ*_*H*_) and *σ*_*H*_~*half N*(0,1). We model the relationship between survival in bioassays with that in an EHT conducted within the same study using:

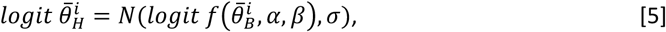

Where *σ* > 0 effectively accounts for idiosyncratic variation about the relationship between bioassay survival and EHT survival. The function *f*(.) captures the mean relationship between bioassay survival and hut survival, and we consider two potential forms for this. A logistic growth model,

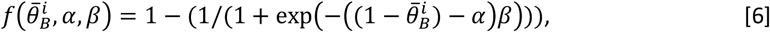

And a log-logistic curve model,

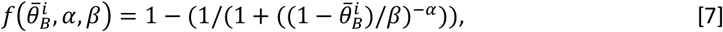

The parameters *σ, α* and *β* are set the following priors: *σ* ~ *N* (0,2), *α* ~ *N* (0,2) and *β* ~ *C* (0,1).

### Association between different EHT outcomes (A2)

ITNs are thought to induce different types of mosquito behaviour change which influence malaria transmission^33^. Following previous work, we investigate the relationship between mosquito survival in EHTs and other factors that influence ITN efficacy to allow the full impact of ITNs to be characterised^20^. EHTs investigating all types of ITN that provided count data for mosquito mortality, blood-feeding and the total number collected in huts were identified. These factors were often estimated across different time horizons, be it daily, weekly or across the whole study (which normally involves a full rotation of all interventions and volunteers sleeping in all huts^13^). The majority of studies examined only report aggregate measures across the full study rather than broken down by day or week. Trials not providing the total count of mosquitoes caught and only reporting the proportion of fed mosquitoes were included in the analyses assuming that the total count in the trial was half the overall median (this aimed to lessen their effect due to uncertainty in the number tested). Data for 24-hour delayed mortality was used, unless the net incorporated the insecticide chlorfenapyr, in which case 72-hour delayed mortality was used to be consistent with its mode of action. Models with all net types grouped together are statistically compared to the relationships for ITNs of different classes. Differences between products within the same class are not currently considered.

#### Deterrence

Volatile compounds from insecticidal interventions may dissuade mosquitoes from entering houses where the intervention has been deployed^32^. This mosquito avoidance behaviour may protect people in houses with interventions but could reduce the overall community effect of indoor vector control tools^33^. Deterrence is defined as the reduction in mosquito entry into houses with an intervention compared to houses without the insecticide^9^. It is typically measured in EHTs by comparing the number of mosquitoes caught in control and intervention huts. The extent to which mosquitoes are deterred by pyrethroid insecticide is thought to diminish in mosquito populations with higher levels of pyrethroid resistance^20,34,35^.

Some EHT studies evaluating ITNs have found higher numbers of mosquitoes caught in intervention huts versus control huts^25,36^. The reason for this apparent “attraction” of mosquitoes to huts containing pyrethroid insecticide is unclear. Different hypotheses for the phenomenon have been proposed though all require further experimental testing with wild mosquitoes^34^. One explanation is more mosquitoes are caught in the intervention arms because they escape the control hut at a greater rate. The window-slit entrances of West African huts are intended to greatly limit mosquito escape back through hut entrances and newer East African huts have been modified to include eave baffles for the same purpose, but escape is likely still possible despite these measures and the exit traps which may be further exacerbated due to differences in collector efficiency. Exit rates may also be lower in intervention huts because the insecticide either kills the mosquito (so it can be counted) or exerts a sub-lethal effect on a mosquito, resulting in it being knocked down or simply disorientated^37–39^. As resistance increases, the combination of fewer mosquitoes being deterred from entering and the sub-lethal effects of insecticide on those individuals which enter and attempt to bite may result in higher counts in intervention huts.

It is not possible to directly estimate the number of mosquitoes escaping control and ITN huts using data that’s available to us. One reason for this is that due to evolution in the designs there is variation in the presence, and style, of baffles between different EHTs over the 15 years of data included in our analysis. Hut designs have been refined to reflect advances in knowledge, though these differences may not be documented. Instead, we use a modelling framework to estimate the relative difference in the probabilities of being caught in the control huts and ITN huts.

We modelled the number of mosquitoes caught in control huts 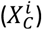 and caught in intervention huts 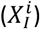 for each trial arm *i* using negative binomial models:

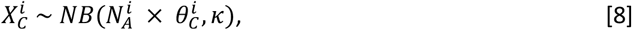

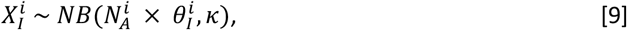

where 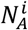 is the number of ambient mosquitoes (immediately outside of the hut), 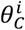 represents the probability of an ambient mosquito entering and being subsequently collected in the control hut and 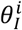 represents the probability of an ambient mosquito entering and being collected in the intervention hut. In eqs. [8] and [9], we use the parameterisation of the negative binomial such that *X* ~ *NB*(*μ, k*) indicates *X* has mean *μ* and variance *μ* + *μ*^2^/*k*, where *k* is an overdispersion parameter. We set the parameters 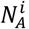, *k* and 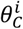 the following priors: 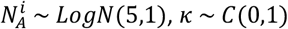 and 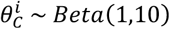

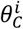 is assumed to remain constant irrespective of the level of EHT survival, whereas 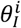 corresponds to the probability an ambient mosquito enters and is collected from an intervention hut given that they were undeterred by the ITN, which is assumed to change with the level of EHT survival.

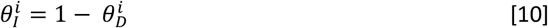

The probability of mosquitoes being deterred 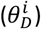 from an intervention hut is assumed to be explained using the following flexible function *g*(.), where parameters *δ*_1_-*δ*_2_ determine the shape of the relationship:

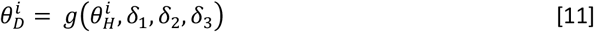

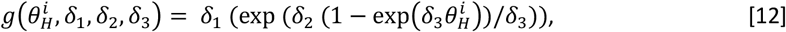

The parameters *δ*_1_-*δ*_3_ are set the following priors: *δ*_1_ ~ *N* (0.8,1), *δ*_2_ ~ *N*(0.8,1) and *δ*_3_ ~ *N*(3,2). We also considered a simple linear function to describe this relationship, but this was ultimately rejected (Supplementary Table 6 and Supplementary Figure 5).

To estimate the difference in the proportion of mosquitoes collected in control huts relative to intervention huts as EHT survival increases, we use the following ratio (*δ*_*C*_):

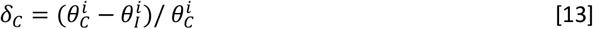

#### Successfully blood-feeding

We define successful blood-feeding as the number of female mosquitoes that blood-feed and survive after entering an experimental hut. All studies which did not include the standard six deliberately made holes in the ITN were excluded from this section of the analyses. For most studies, it was not possible to determine whether mosquitoes had fed and survived or fed and died as aggregated feeding data are presented. As in Sherrard-Smith et al.^21^, we adjusted the number of fed mosquitoes (*N*_F_) by the proportion that died, which implicitly assumes that feeding and dying are independent events. *N*_*H*_ is the total number of mosquitoes collected in the hut and *N*_D_ is the total number of mosquitoes that died.

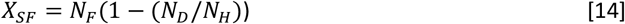

We modelled the number of mosquitoes successfully blood-fed in each trial arm 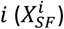 as a binomial model:

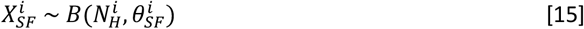

where 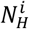 is the total number of mosquitoes collected in the hut and 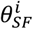 represents the probability of successfully blood-feeding, which is itself modelled:

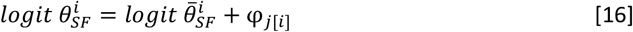

where, as above, φ_*j*[*i*]_ is a location-level effect corresponding to the location, *j*, where trial arm *i* took place. Here, 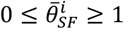 represents the estimate of successful blood-feeding in the trial after accounting for location-level variability. As before, we set the following priors: φ_*j*[*i*]_~*N*(0, *θ*) and *σ*~*half N*(0,1). We model the relationship between successful blood-feeding and survival in the same hut trial using:

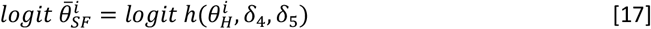

where *σ* > 0 effectively accounts for idiosyncratic variation about the relationship between hut trial survival and successful blood-feeding. The function *h*(.) captures the mean relationship between hut survival and successful blood-feeding:

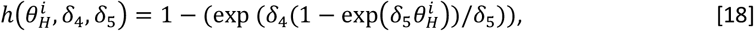

where the parameters are set the following priors: *δ*_4_~N (0,0.01) and *δ*_5_~N(8,0.3)

#### Exiting unfed

Those mosquitoes which enter a hut and do not die or successfully feed, are assumed to exit the hut without feeding. Mosquitoes are defined as having exited irrespective of whether they were caught in exit traps or collected (unfed and alive) from within the hut. Therefore, the probability of a mosquito exiting unfed 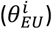 in trial arm *i* can be estimated by 1 minus the probability of successfully blood-feeding and the probability of dying:

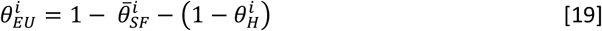

Bayesian models were fit to the data using a version of dynamic Hamiltonian Monte Carlo sampling implemented via Stan^40^ and can be found in the supplementary material. Four chains were run for 4000 iterations each, with the first 2000 iterations discarded as warm-up. The posterior distributions and 95% Bayesian credible intervals were estimated, with the posterior predictive fits of the model to data shown throughout the text (e.g. Fig. 3B, C & D).

**Figure 1.**
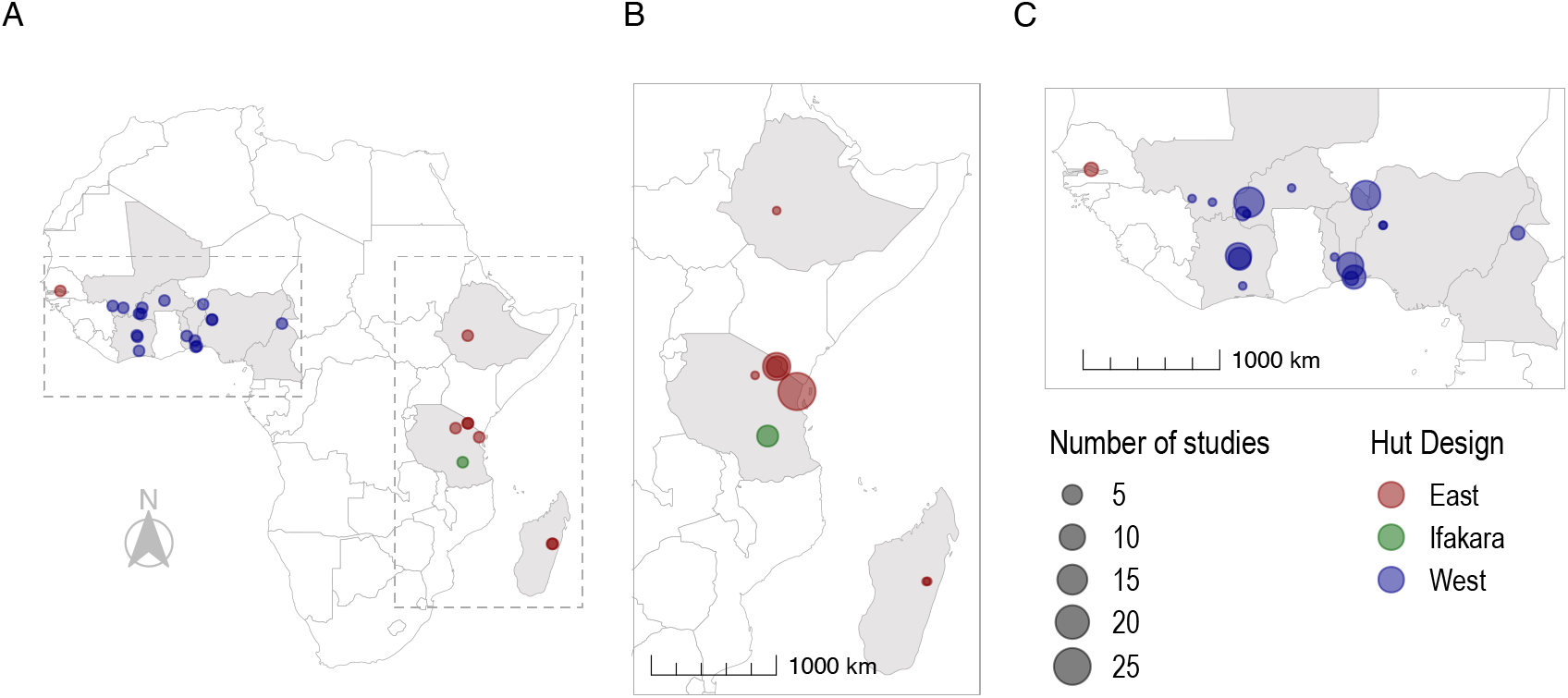
Maps of EHT sites where ITNs and/or IRS have been investigated. Panels show sites across (A) Sub-Saharan Africa; (B) Eastern Africa (54 trials), including Ethiopia, Tanzania and Madagascar; (C) West (80 trials) and Central Africa (1 trial), including The Gambia, Mali, Ivory Coast, Burkina Faso, Togo, Benin, Nigeria and Cameroon. Point size indicates the number of studies at each site, whilst point colour denotes the design of hut used, be it East African (red), Ifakara (green) or West African (blue).

**Figure 2.**
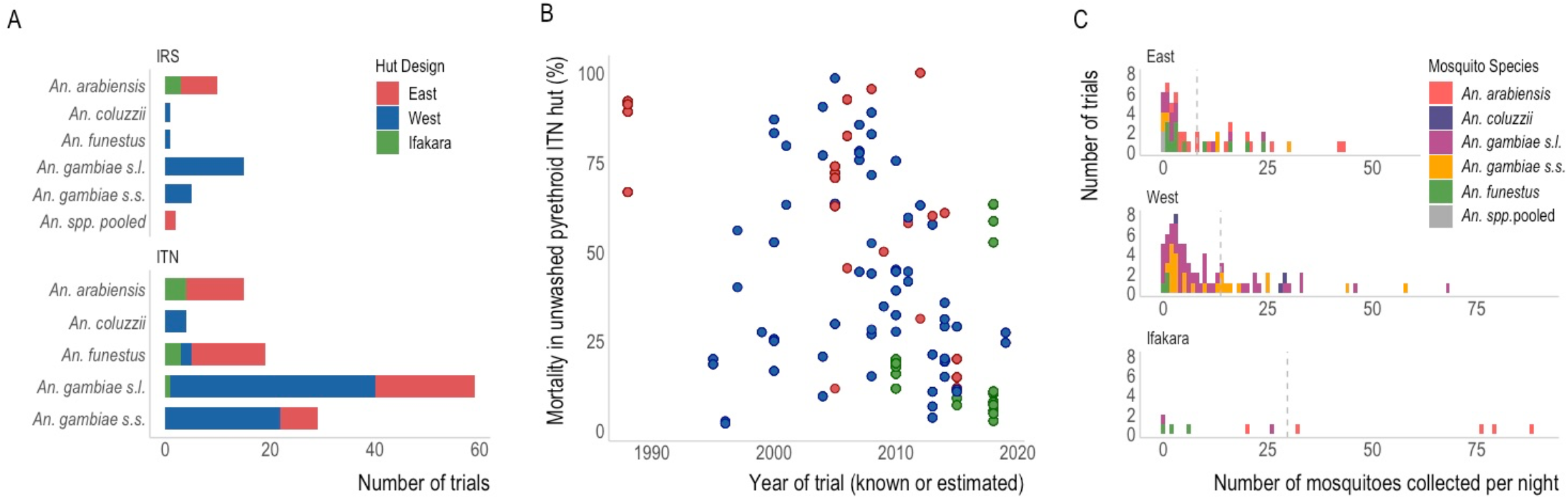
Characteristics of EHT data. A) The number of EHTs investigating IRS or ITNs that collected data on each hut design and mosquito species (as reported in the study). Bar colours indicate the design of the hut used in the trial as shown in the legend, be it East African (red), West African (blue) or Ifakara (green). B) Summary of the reported mortality observed in hut trials evaluating unwashed pyrethroid-only ITNs and how these change over time. Points are coloured according to the hut design (see A). C) Histograms showing the average number of mosquitoes collected per night in the control huts of EHTs. The dotted line represents the mean number collected in trials of each hut design. Note: Some trials collected multiple mosquito species, therefore single trials may be counted more than once in this figure.

**Figure 3.**
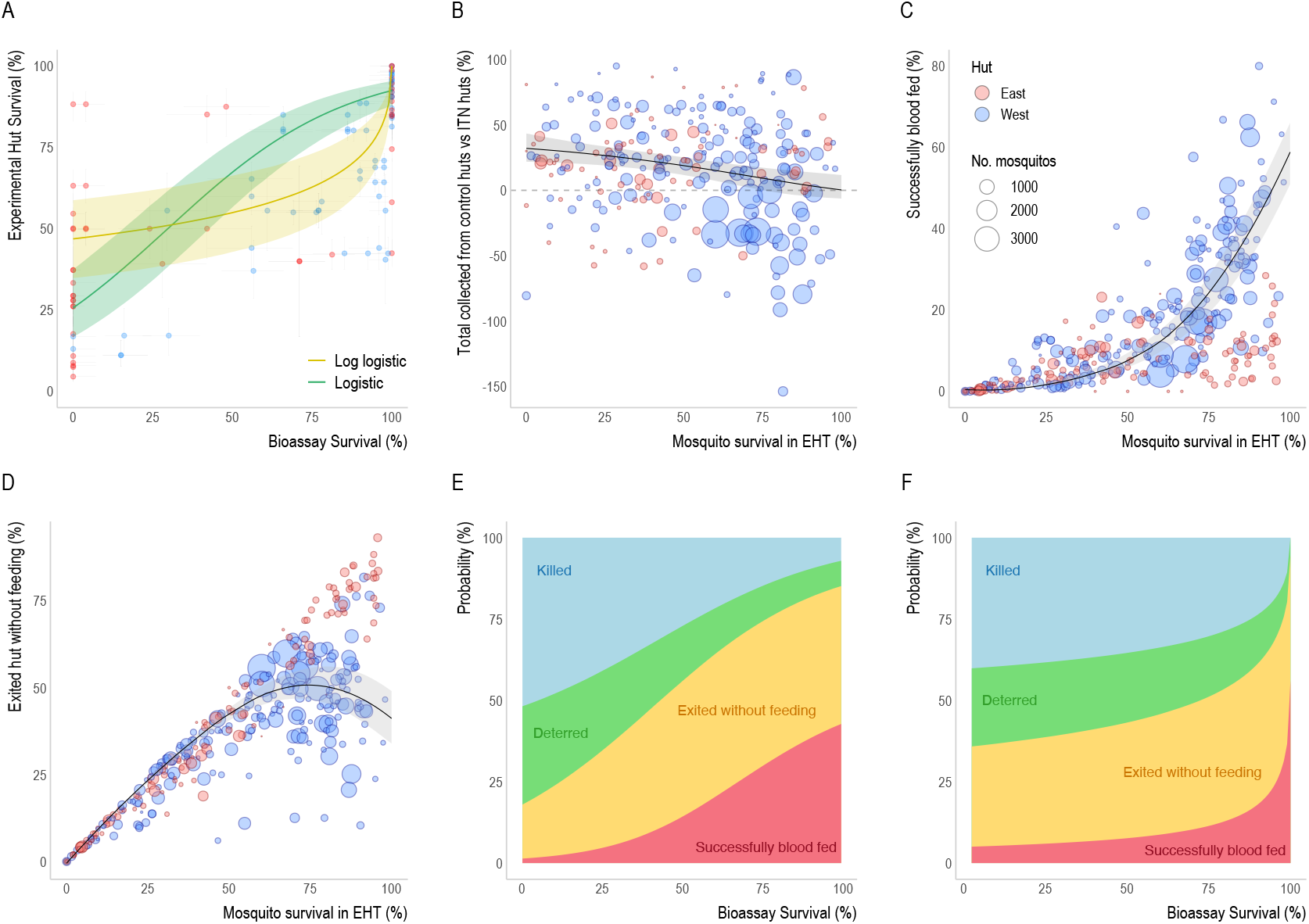
Entomological outcomes as predicted by bioassay survival. A) Comparison of the level of pyrethroid resistance as measured using a discriminating dose bioassay and the percentage of mosquitoes which enter a hut with a pyrethroid-only ITN and survive 24 hours after being collected. Solid lines show the fitted relationship using either the logistic (green) or log-logistic (yellow) models. Vertical and horizontal lines show 95% confidence interval estimates for each data point. B-C) show the relationship between EHT survival (24h post-collection, unless the ITN incorporated the insecticide chlorfenapyr, in which case 72h survival was used) and the probability of B) being caught inside a control hut relative to a hut with any type of ITN (where anywhere above the grey dashed-line indicates more mosquitoes were caught in the control hut), C) mosquitoes successfully feeding and surviving, or D) exiting the hut without feeding. The point size in B-D is proportional to the total number of mosquitoes collected in the trial and coloured according to hut design: East (red) or West (blue). Solid lines in A-D show the best fit relationship whilst the lighter shaded area indicates 95% credible intervals for the best fit curves. E-F) The models from A-D were combined to summarise how the average probability that blood-feeding mosquitoes will be killed, exit without feeding, deterred from entering or successfully blood-feed, varies with bioassay survival. The relationship between bioassay and EHT survival is highly uncertain so both the logistic model (E) and the log logistic model (F) are presented.

#### Outcomes of a feeding attempt as resistance increases

In a feeding attempt, a mosquito is either deterred from entering a hut by the ITN or is undeterred and enters the hut, where they then either die, successfully blood-feed or exit unfed. To show the relationship between these outcomes and resistance, EHT survival was translated to bioassay survival using function [6] and [7].

## Results

### Summary of EHTs in Sub-Saharan Africa

The review identified 115 eligible studies reporting data from 135 experimental hut trials (EHTs) (Supplementary Figure 1). These trials were conducted at 26 different sites across 11 countries in Sub-Saharan Africa. The majority of these sites were located in western Africa (n=17), with only 1 site in central Africa and 8 across eastern Africa (Figure 1). Muheza, Tanzania was identified as the site with the highest number of trials (n=26), followed by Kou Valley, Burkina Faso (n=15) and Malanville, Benin (n=14). Most trials were carried out in western Africa (n=80), whilst 54 trials were performed in eastern Africa and 1 in central Africa.

The earliest trial identified used the East African hut design and took place in 1988, whereas the first trials identified using the West African and Ifakara designs were in 1995 and 2010 respectively (Supplementary Figure 2A). The greatest number of trials identified in the analyses of the literature were conducted in 2008 (n=16) with high numbers reported between 2010-2015 (Supplementary Figure 2A).

Of the 135 trials identified, 58% were carried out in the West African hut design (n=79), 37% used East African huts (n=50) and just 4% of trials used Ifakara huts (n=6). The majority included at least one intervention arm that investigated ITNs (n=112). There were far fewer trials investigating IRS (n=33) and only 6 trials were identified that looked at ITNs used in combination with IRS. There were a small number of ITN or IRS trials that included EHT arms exploring the use of other intervention types (n=7), which include insecticide treated plastic sheeting, wall linings, net wall hangings and coils.

In Figure 2A, we summarise the EHT data according to the collected species in trials involving IRS (top) and ITNs (bottom). The most frequently reported mosquito species in the study area of trials was *Anopheles gambiae* sensu lato (*s.l*.) (n=128, 85%). Only 46% of the trials which identified members of the *An. gambiae s.l*. complex identified mosquitoes to the species level, including *Anopheles arabiensis* (n=22, 17%), *Anopheles coluzzii* (n=5, 4%) and *Anopheles gambiae* sensu stricto (*s.s*.) (n=32, 25%). There were 20 trials where *Anopheles funestus* was present (13%) and 2 trials where the results were presented for all Anopheles species pooled together. A total of 15 trials identified more than one mosquito species during a trial, though this may be because many studies aggregated data to the level of the species complex. Of these 15 trials, 11 reported total counts of mosquitoes and the dominant mosquito species comprised on average 67% of all mosquitoes caught (range 52-98%).

Overall, there is a decline in mortality seen in EHTs evaluating unwashed pyrethroid-only ITNs over the last 20 years (Figure 2B). During this time the level of mortality observed in trial arms with untreated nets remained broadly constant indicating pyrethroid-only ITNs are now killing fewer mosquitoes entering experimental huts. Similar reductions in blood-feeding inhibition induced by pyrethroid-only ITNs were also observed over time (Supplementary Figure 3, Sup. Table 5).

The mean number of mosquitoes collected per night in control huts of each hut design were n=9 for East, n=12 for West and n=33 for Ifakara huts (Figure 2B). Across all designs, there was considerable variability in the number of mosquitoes caught in control huts per night, ranging from 0 to 43 for East, 0 to 74 for West and 0 to 88 for Ifakara huts (Supplementary Figure 2B). There was a strong association between the species of mosquitoes caught and hut design, likely reflecting the mosquito species composition in the sites that the different huts were built (Figure 1). As a result of this, our analyses could not disentangle differences in behaviour across species versus that induced by hut design. For example, *An. arabiensis* was only caught in an East African or Ifakara hut design trial whilst *An. coluzzii* was only identified in West African huts (Figure 2A). *An. funestus* mosquitoes were identified in sites of different hut designs, though the numbers reported in the West African huts were very low (fewer than 2 mosquitoes per night). Consequentially, although this is sub-optimal, all future analyses group all species together to examine differences in hut design. As such, care should be taken when interpreting our results, since the entomological impact of ITNs likely reflects differences in hut design and local factors associated with mosquito species (and other unmeasured factors).

Analyses are restricted to the East and West African hut designs as the number of trials identified using huts of the Ifakara style are relatively low, they were only carried out in one trial site, and the data appeared to exhibit outlying trends (Figure 2B, Supplementary Figure 4).

### Pyrethroid resistance and ITN mortality

Thirty-seven EHTs were identified where concurrent pyrethroid discriminating dose bioassays and EHTs of pyrethroid-only ITNs were conducted on the same mosquito population (Supplementary Table 3).^41–71^ After excluding Ifakara huts, there were 107 datapoints (paired bioassay survival and EHT survival) from 34 EHTs. The higher the reported bioassay resistance level in a mosquito population, the more likely mosquitoes were to survive 24 hours after entering a hut with a pyrethroid-only ITN (Figure 3A). The shape of the relationship is highly uncertain, with no clear pattern showing how EHT survival changes with increasing levels of resistance. Despite there being a clear positive association between bioassay and EHT survival, there was considerable variability in the data. To reflect this uncertainty, we fit two functions representing the mean association between bioassay survival and EHT survival to the data – logistic and log-logistic models. The logistic model suggests 25% of mosquitoes survive in EHTs when mosquito populations are deemed fully susceptible to pyrethroids (i.e. 0% bioassay survival). Conversely, the log-logistic model indicates there is greater EHT survival (~50%) in a fully susceptible population, with incremental changes in reported bioassay survival being less predictive of changes in EHT survival until very high levels of resistance. This more gradual reduction in ITN-induced mortality with increasing resistance suggests the impact of pyrethroid resistance at low to moderate levels may be less. Both models fit the data equally well (leave-one-out cross-validation indicates that there was a negligible difference between the models, with an expected log predictive density (elpd) of −844.9 for the logistic versus −846.6 for the log-logistic model (higher values indicating better fit)). Similar trends are observed if the same functions are fit to trials of different hut design (Figure 4A). Twenty-four-hour mortality measured in Ifakara huts appeared consistently low regardless of the level of susceptibility to pyrethroids, further justifying their exclusion from these analyses (Supplementary Figure 4).

**Figure 4.**
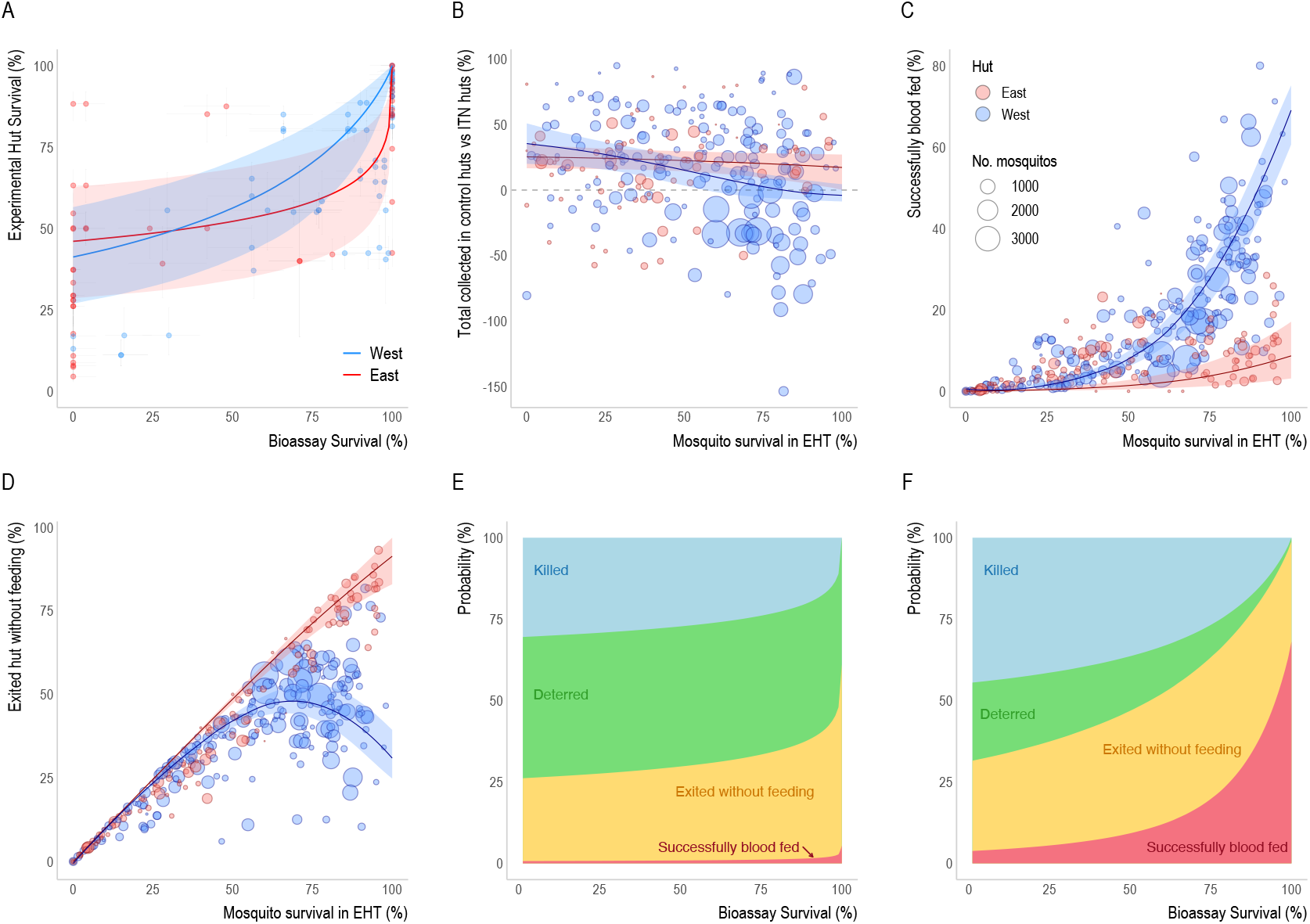
Entomological outcomes as predicted by bioassay survival according to East and West African hut designs. A) Comparison of the level of pyrethroid resistance as measured using a discriminating dose bioassay and the percentage of mosquitoes which enter a hut and survive 24 hours. Solid lines show the fitted relationship for the East (red) and West (blue) African hut design assuming the log-logistic function. Comparable plots showing the logistic function are provided in Supplementary Figure 7A. B-C) show the relationship between EHT survival (24h post-collection, unless the ITN incorporated the insecticide chlorfenapyr, in which case 72h survival was used) and the probability of B) being caught inside a control hut relative to a hut with any type of ITN (where anywhere above the grey dashed-line indicates more mosquitoes were caught in the control hut), C) mosquitoes successfully feeding and surviving, or D) exiting the hut without feeding. The point size in B-D is proportional to the total number of mosquitoes collected in the trial and coloured according to hut design. Solid lines in A-D show the best fit relationship whilst the lighter shaded area indicates 95% credible intervals for the best fit curves. E-F) The models from A-D were combined to summarise how the average probability that blood-feeding mosquitoes will be killed, exit without feeding, deterred from entering or successfully blood-feed, varies with bioassay survival for either East (E) or West (F) African hut design. Both (E-F) show the log-logistic model, see Supplementary Figure 7B-C for models using the logistic function.

### Mosquito response to ITNs

Eighty-two EHTs investigating ITNs of all types were included in the analyses comparing different EHT outcomes (Supplementary Table 4)^46–117^. In addition to mortality data, 65 of these provided the total number of mosquitoes collected and 81 provided data for blood-feeding. The following section reports models fit to data for all ITNs, irrespective of the insecticide used, as comparisons with models restricting data to pyrethroid-only, pyrethroid-PBO or pyrethroid-combination nets showed no significant improvement in fit (Supplementary Table 7).

On average, fewer mosquitoes were collected in huts with ITNs compared to control huts, but as the level of survival in EHTs increased, the difference between the number of mosquitoes collected from control versus ITN huts fell (Figure 3B). For example, in susceptible mosquito populations (where <25% of mosquitoes that enter ITN huts survive), on average 20% more mosquitoes were collected in control huts. Whereas, once all mosquitoes survive, the number of mosquitoes collected from control huts is estimated to be equal to the number in ITN huts (Figure 3B). This suggests that the level of ITN-induced deterrence appears to decrease in mosquito populations with higher levels of pyrethroid resistance. Similar patterns were seen in huts of both East and West African design (Figure 4B). In both cases, the difference in the number of mosquitoes collected in control huts versus intervention huts declined, but this trend was much more evident in data from West African huts. Once mosquito survival increases beyond 85%, the model indicates the number of mosquitoes collected from West African ITN huts exceed the number in control huts. Fitting the models across data from each hut design separately provided a significantly better fit than when pooling analyses (Supplementary Table 6). Fitting an alternative linear relationship resulted in slightly poorer fits than the non-linear model in both pooled and hut-design specific analyses (Supplementary Table 6 & Sup. Figure 5). As part of deriving the relationship shown in Figure 3B, we also estimate the probability of deterrence and the probability of escape (i.e. exiting the huts without being caught in exit traps) from the control hut, and these are shown in Supplementary Figures 5 & 6. The model suggests that there are greater numbers of mosquitoes escaping from the West African design of hut. This is driven by a greater number of studies reporting more mosquitoes in ITN huts compared to control huts at high levels of pyrethroid resistance (Figure 4B).

The proportion of mosquitoes that successfully blood-feed (feed and survive) increases dramatically with survival, with the model predicting on average around 60% of mosquitoes successfully blood-feeding at 100% survival (Figure 3C). There is a substantial difference in the proportion of mosquitoes that successfully blood-feed as survival increases between the different hut designs (Figure 4C). There is a more dramatic rise in West African huts, with over 60% of mosquitoes estimated to successfully blood-feed at 100% survival. In contrast, less than 10% of mosquitoes are estimated to successfully blood-feed in East African huts when all mosquitoes survive. If a mosquito does not die or successfully blood-feed, it is assumed they will ultimately exit the hut without feeding. This means that the proportion of mosquitoes exiting unfed decreases at high mosquito survival due to increases in blood-feeding, especially in West African huts (Figure 4D).

We can use the estimated relationships shown in Figure 3A-D to predict what happens to a mosquito when it attempts to blood-feed on a person sleeping under an ITN. Due to the dependency on bioassay survival for each of these relationships, the likely outcome of a feeding attempt depends on the local pyrethroid resistance level. In Figure 3E-F, we show the probability of death, deterrence, exiting without feeding and successfully blood-feeding as a function of bioassay survival: panel E shows the results for the logistic model and panel F shows the log-logistic. 3E shows gradual changes in ITN efficacy as resistance rises, with ITN-induced killing and deterrence falling to less than 10% and successful blood-feeding estimated to increase from just over 1% to 42%. By contrast, when the relationship between hut survival and bioassay survival is described by the log-logistic fit (Figure 3F) there is a much sharper change in entomological outcomes at very high resistance levels. Successful blood-feeding is estimated to increase even further, from 5% to 56%, whilst killing and deterrence reduced to 0% and 5% respectively. The estimates for the outcome of a mosquito feeding attempt using the log-logistic fit for the East and West African hut design are shown in Figure 4E and 4F respectively, with plots using the logistic fit in Supplementary Figure 7.

## Discussion

As pyrethroid resistance rapidly spreads across sub-Saharan Africa, it is important to understand how the entomological impact of vector control interventions are changing so the most effective malaria control tools can be deployed. Experimental hut trials (EHTs) are an affordable and relatively quick method for determining the efficacy of interventions that target the mosquito within the home. They were originally developed to assess the impact of insecticidal interventions which mostly delivered fast-acting neurotoxic insecticides, relative to a control arm without the insecticide. The rise of pyrethroid resistant mosquitoes and the development of alternative classes of ITNs and IRS means that they are increasingly being used to assess the efficacy of different interventions and compare products. This shifting of the question from “does something work” to “how well does it work” requires a better understanding of the measurement error of this complicated real-world assay, particularly as alternative classes of ITN may have slow-killing effects, or sublethal effects that are not well demonstrated in an EHT. Evaluation of the impact of pyrethroid resistant mosquitoes necessitates comparison between sites with different mosquito populations so results need to be compared between different trials. However, differences in the design of hut used in different sites make it hard to directly compare EHT results. This systematic review compiles all published data and known grey literature to investigate the differences between West African and East African hut designs and more accurately quantify the impact of resistance on the entomological efficacy of ITNs.

The review identified 135 experimental hut trials characterising the entomological efficacy of ITNs and IRS. The number of trials conducted appears to be increasing over time, reflecting the importance of this assay in evaluating new indoor vector control tools. Fewer studies were identified reporting data collected after 2016 (Supplementary Figure 2A) likely reflecting the delay between the trial and publication of results rather than a drop in the number of EHTs conducted. The trials evaluating pyrethroid-only ITNs were conducted during a time when local mosquito populations exhibited a range of susceptibility to pyrethroid insecticides. A decline in EHT mortality and blood-feeding inhibition induced by pyrethroid-only ITNs is seen over time (Figure 2B, Supplementary Figure 3B, Sup. Table 5) which is likely to result from changes in the underlying susceptibility of the local mosquito population seen over the same time-period^10^. These trends provide strong evidence of the diminished entomological efficacy of ITNs since the year 2000, though other explanations are also possible. This study groups together all ITNs and earlier studies are more likely to be dipped in insecticide (CTNs) rather than those which incorporate or impregnate the insecticide into the netting (LLINs). Differences in the dominant vector species, subtle changes in hut design and study protocol, and the use of other interventions targeting mosquitoes in the region, are likely to influence results, though there were insufficient data available throughout the time-period to investigate these potentially confounding factors.

The reduction in pyrethroid ITN mortality over time is further supported by the association between bioassay and hut trial survival observed in studies which reported both assays conducted on the same mosquito population (Figure 3A). Unfortunately, only a minority of EHT studies (37/135) reported bioassay data in the same publication. This lack of data makes the shape of the relationship between these metrics highly uncertain. The unclear association between the more controlled pyrethroid bioassay and the more real-life EHT is to be expected, though it is likely to be exacerbated by the measurement error in both. The discriminating dose bioassay is known to be highly variable, with the accuracy depending on the ambient conditions under which the assay is conducted, the type of assay used, and the number of mosquitoes exposed (which in some instances appears relatively low)^31^. EHTs tend to have data from many more mosquitoes (when summed across the length of the trial), but also have considerable variability, reflecting the greater biological realism of EHTs. Mosquitoes sampled and used in the bioassay will be different from those which naturally enter experimental huts in their species composition, age and factors related to their rearing. For example, bioassays which use mosquitoes reared from larvae may be collected from a small number of breeding sites at a single time point, whilst EHTs report data from mosquitoes which are likely to have developed in a more diverse set of breeding sites over the period of multiple months. The relationship between bioassay and EHT survival has been used to infer how the effectiveness of ITNs changes with the level of pyrethroid resistance^20^. Given the variability in these data we used two different functional forms to characterise the relationship. Fits for the logistic model indicate a more linear relationship between bioassay and EHT survival, which is relatively similar to previous models fit with smaller datasets, though the gradient of the change is more gradual^20^. Fits with the log-logistic model suggest that survival in EHTs with pyrethroid ITNs is higher in susceptible mosquito populations, though changes less as bioassay survival increases until nearly all mosquitoes survive exposure in the bioassay. This model suggests the greatest difference in hut trial survival is seen when between 80-100% of mosquitoes survive exposure in a bioassay. The shallower gradient of the relationship may have important implications for understanding how resistance may change over time. It suggests that the loss of efficacy of pyrethroid ITNs could be more gradual than the changes in mortality observed in the discriminating dose bioassays. Further work is needed to understand if this is true or an artifact of the assay used for approximating resistance. Discriminating dose bioassays were originally designed to identify resistance when it first appeared and were not intended to compare populations with higher levels of resistance, where assays such as dose response (intensity) assays are recommended^27^. Susceptibility and tube bioassays expose mosquitoes to a lower concentration of insecticide than is typically used on ITNs, so may not be able to distinguish between populations with moderate and high EHT survival. Grouping these data together would tend to make the relationship between bioassay and hut trial survival steeper in more resistant populations, which may not be the case if other tests were used. It is important to realise that changes in the shape of the relationship translating field measurements of resistance (measured using the discriminating dose bioassay) to ITN entomological efficacy (measured in the EHT) does not alter the public health impact of resistance which has shown to be substantial^118,119^. It does however highlight why observational studies looking for associations between bioassay data and malaria may have been inconclusive^120^, as large differences in bioassay survival may not always translate into substantial changes in ITN efficacy. This will have important implications for how ITN epidemiological efficacy might be predicted to change in a site over time and what methods we should use to track susceptibility to insecticides.

To try to understand how mosquito behaviour changes with susceptibility, all ITN data were analysed together, grouping together nets with different insecticides (with and without synergists) without separating trials using East and West African huts. There is a strong association between survival of mosquitoes which enter ITN huts and the probability of mosquitoes being deterred, successfully blood-feeding and exiting without feeding, though there is considerable variability between studies - reflected by point estimates often falling a long way from the best fit line. The ratio between the number of mosquitoes collected in the control huts compared to ITN huts is particularly variable, with some huts exhibiting high survival having twice as many mosquitoes collected in the control hut, whereas some have twice as many mosquitoes in the ITN hut (Figure 3B). On average, the model predicts deterrence is high in susceptible mosquito populations and falls to zero as survival increases.

Here we assume that no mosquito escapes from huts with ITNs and the level of deterrence may be greater if this is not the case. Our EHT data were unable to directly estimate mosquito entrance and escape so it is unclear why more mosquitoes are often collected in control huts compared to intervention huts. The deterrent effect of ITNs and how it changes with resistance is thought to be an important component of the effectiveness of indoor vector control,^24,121^ so further work is needed to verify this behaviour and evaluate other hypotheses for the possible cause^36^.

The probability of mosquitoes feeding and surviving - termed ‘successful blood-feeding’ - rose exponentially with increasing survival. When all mosquitoes that enter a hut survive, the model predicts that those mosquitoes have a probability of successfully feeding of almost 60% (Figure 3C). This high loss of blood-feeding inhibition is broadly consistent with a previous study^20^, though the current systematic review contains recent studies with higher levels of mosquito survival, exacerbating the trends. The non-linear increase in blood-feeding with increased survival indicates ITNs provide personal protection until high levels of survival, which is likely to have important implications for the transmission of malaria. Those mosquitoes that do not blood-feed or die are classified as having exited the hut unfed. It is unclear whether these mosquitoes are able to attempt to re-feed immediately or whether they may experience some sub-lethal impact of the insecticide^122–124.^

Experimental huts were originally designed so that their structure reflect the type of houses found within the region. This study has shown that most EHTs have been carried out in Western Africa, reflected by the greater number of trials conducted with West African hut design. Studies in East Africa are carried out in either East African or Ifakara huts, with the latter being a relatively new design developed to more accurately represent the houses in villages in south eastern Tanzania^17^. This work provides strong evidence that hut design influences hut trial outcomes. This is consistent with the two previous small EHTs^18,19^ that directly compared hut design in the same site, unlike this study, which examines differences between sites. Ifakara huts are much larger, with a volume of 51.35m^3^ compared to 18.90m^3^ and 13.3m^3^ for East and West African huts respectively^18^. The mean number of mosquitoes collected per night in Ifakara huts is over double the mean collected in either East or West huts. Whilst this could reflect location differences in mosquito densities or species, when directly compared, Massue et al found that Ifakara huts collected 3 to 4 times the number of mosquitoes than the other designs.^18^ Greater mosquito collections are beneficial in increasing the power of EHTs, but it is possible that the size of the hut has contributed to the much lower mosquito mortality observed, regardless of the level of resistance measured in the mosquito population. In a larger hut, mosquitoes could be less likely to come into contact with a net and therefore the interventions may appear to induce lower mortality. This finding is in contrast with a Benin study, which found greater mortality in Ifakara huts compared to West African huts, though these findings were based on nuisance Culex mosquitoes as opposed to anopheline mosquitoes^19^. The lack of a clear picture illustrates the difficulty in disentangling the impact of hut design and the response of the local vector population that enter these huts. Due to this stark difference in mortality (Supplementary Figure 4) and the very few Ifakara trials identified, Ifakara huts were not included in the probability predictions for the outcomes of a mosquito feeding attempt.

The relationship between bioassay and hut trial survival appears substantially different for East and West African huts, with, on average, higher survival seen in West African huts for a given level of bioassay mortality. Care should be taken in interpreting this result due to both the difference in mosquito species between EHT sites and because of the sparsity of data which makes the shape of the relationship highly uncertain (differences in the logistic curve presented in Figure S7 suggest the difference is less assuming this functional form). There were more distinct differences in the estimated relationships between hut trial survival and other mosquito behaviours for EHTs with East and West African designs. The probability of deterrence from ITN huts, and therefore the ratio between the probability of mosquitoes being collected in control huts compared to ITN huts, fell more sharply in West African huts than East African huts. Beyond 80% survival, slightly more mosquitoes were collected from West African ITN huts than control huts. Similar trends have reportedly been observed in Ifakara trials too. It is unclear why data from East African huts does not exhibit the same trend, as higher numbers of mosquitoes are always caught in control huts of this design. East and West African huts differ in their size, eaves, window size and the use of baffles, though differentiating the difference of these design factors is beyond the scope of these data. Overall, more mosquitoes were caught in control huts with a West African design than an East African design, though this is likely to result from differences in productivity in the breeding sites nearby than solely through differences in mosquito escape. It is important to note that there are relatively few datapoints at higher levels of survival for East African huts, so this difference in the relationship should be interpreted with caution.

The greatest disparity between hut designs was in the relationship between hut trial survival and successful blood-feeding. The probability of mosquitoes entering a hut and successfully feeding in West African huts increased exponentially to over 60%, whilst it never exceeded 10% in East African huts (Figure 4C). *An. arabiensis* mosquitoes often found in East Africa have been shown to have a lower propensity to blood-feed on humans than other Anopheles species and are also more likely to bite outdoors.^28–32^ From visual inspection of the data, this low-level blood-feeding did not appear to be specific to a particular mosquito species. Nevertheless, differences are likely to exist, so comparison of East and West African hut results should stratify by mosquito species. It could also be suggested that due to the smaller size of West African huts, mosquitoes may be more likely to come into contact with the volunteer sleeper and feed. Mosquitoes evaluated in the West African hut design are also able to move unimpeded between the veranda exit trap and the hut, potentially having more opportunities to feed. In contrast, mosquitoes in East African huts are channelled in one direction by air flow, potentially making them less likely to return into the hut. Where data are aggregated, we have made an assumption of independence in those that feed and die or feed and survive. Further work is needed to understand the behaviour of mosquitoes in huts of different design, potentially using video tracking technology to directly observe feeding outcomes of individual mosquitoes^130^.

The statistical models allowed the impact of insecticide susceptibility on the efficacy of ITNs to be summarised as the relationship between bioassay survival and the outcome of a single mosquito blood-feeding attempt. These summaries can be used to parameterise mathematical models of the transmission dynamics of malaria^20^ to estimate the public health impact of ITNs. The uncertainty in the bioassay and EHT survival relationship means that there is still some ambiguity in the rate at which pyrethroid ITN efficacy changes with increasing resistance. Both models suggest that low levels of resistance reduce the ability of ITNs to kill mosquitoes and will likely reduce the community impact of mass ITN distribution. Nevertheless, it is important to note that at these moderate levels of resistance the results suggest pyrethroid-only ITNs still provide users with personal protection by reducing blood-feeding even when ITNs have holes. This personal protection is predicted to persist until relatively high levels of resistance. This is especially the case if there is a more gradual change in hut trial survival with increasing pyrethroid resistance (as illustrated by the log-logistic model).

There are also differences in EHT outcomes when the models are fit to all data together or separated by hut type. In East African huts, ITNs are estimated to provide a higher degree of personal protection against moderately resistant mosquito populations, but this protection dramatically declines against highly resistant populations. Which EHT design best represents the efficacy of ITNs in the field is unclear. Models parameterised with each of these options could be compared to field data to select the predictive measure. Whatever the result, no hut trial design can adequately represent the diverse array of housing seen across a region. Large scale observational studies have shown housing type does influence malaria prevalence but there was no evidence of changes in the effectiveness of ITNs^131^. Hut design does however influence EHT outcomes so care should be taken when comparing products evaluated in different hut designs, especially when comparing blood-feeding. Comparison of interventions relative to control huts (i.e. induced mortality or blood-feeding inhibition) may reduce these differences, though this may also bias results if absolute values vary substantially. A standard hut design allows differences between local mosquito populations and the intervention to be systematically compared without the confounding factor of hut design. Researchers could therefore consider building huts of an existing design to allow results to be more easily interpreted and compared to other trials, even if these designs do not match the predominant local house type. If this approach is taken it will be important to account for the real-world differences in housing type over time to understand whether they influence the efficacy of indoor vector control interventions.

These analyses have some notable additional limitations. This is predominantly due to sparsity of data which in particular limited differentiation between mosquito species. The majority of trials report results from the *An. gambiae* sensu lato (*s.l*.) species complex and do not distinguish between individual members which have very different behaviour. This means that all mosquito species are considered the same given the level of EHT survival, which is unlikely to be the case. Clearly, it would have been beneficial to be able to differentiate between species that are known to differ in their behaviour. Nevertheless, grouping all species together provides a snapshot of the efficacy of ITNs against indoor-biting mosquitoes during the study period, and is therefore likely to be representative of the impact of ITNs on indoor feeding mosquitoes in that site. It would have been useful to compare different vector control products and how their estimated efficacy varies between studies and hut designs^119,132,133^, but there were too few datapoints for trials using the same net brands to investigate this thoroughly. The vast majority of studies only reported summary data making it impossible to differentiate whether mosquitoes that died had fed or not. We encourage future trials to submit all disaggregated raw data when publishing trial results to allow differences between huts, volunteers and changes over the course of the study to be disentangled and allow more precise efficacy estimates. It would also be valuable for trials to measure retention of mosquitoes within huts as a matter of course to enable us to quantify the escape that is unaccounted for by exit traps. This study has grouped together a wide range of ITNs from different classes, for example, with and without synergist or with an additional active ingredient. Though the level of mortality induced by different ITNs varies, this study shows that the relationship between mortality and other behaviours measured in EHTs are relatively consistent. This should be verified further as the number of EHTs increases and novel ITNs with new modes of action are evaluated. Similarly, we do not differentiate between products within a class of ITNs. This was due to the high between-study variability, which means that products need to be directly compared within the same trial to allow meaningful comparisons. There is currently a sparsity of these studies, so though there may be substantial differences between products, it could not systematically be evaluated here. Finally, the use of discriminating dose bioassays to measure resistance is likely to become increasingly problematic as resistance spreads.

These assays are currently the most widely used method for assessing resistance in field populations but are unable to differentiate between highly resistant populations when dose response assays or the use of genetic markers of resistance will be required. Future EHT studies of new and existing products should be encouraged to characterise the local mosquito population as much as possible using different metrics, such as these measures of resistance, to allow results of EHTs to be extrapolated to different field populations.

This systematic review highlighted gaps in EHT literature. ITNs are the main method of malaria control so the results of EHTs are applied across sub-Saharan Africa and beyond. Despite this, there were only 26 study sites identified in 11 countries where products were evaluated. For example, there were very few studies conducted in central Africa, where only 1 trial was identified in northern Cameroon. Given the substantial financial investment in vector control and the importance of ITNs and IRS in public health it would be beneficial for trials to be carried out in more geographically diverse settings to allow these products to be tested against a wider range of mosquito species. The review also identified that very few trials investigated the use of IRS in combination with ITNs, and none that showed the impact over a long time-period as insecticide decayed on the walls. The added benefit of this combination of vector control tools is unclear, with varying results found in RCTs.^134–136^

## Conclusion

These analyses present EHT predictions for the changes to the entomological efficacy of pyrethroid-only ITNs used against mosquito populations with increasing levels of pyrethroid resistance. The EHT is a hugely important assay which provides high quality data on indoor vector control tools in near real-world settings. Results show the impact of pyrethroid resistance on the interaction between mosquito, human and ITNs is substantial and highly complex. It highlights that EHT results can be variable, and the causes of this variability are often unknown, meaning that results should be interpreted with care. Nevertheless, the high number of trials conducted with diverse populations of mosquitoes across Africa means that this variability can be characterised and accounted for, allowing EHTs to provide unparalleled information on some of the world’s most important disease control interventions.

## Data Availability

Data extracted from published studies will be provided in the supplementary material of the published manuscript. Unpublished data that support the findings of this study will be available from the corresponding author upon reasonable request and subject to agreement with the data owners.

## Acknowledgements

The work was supported by the Innovative Vector Control Consortium (IVCC), the Wellcome Trust [200222/Z/15/Z] MiRA and the UK Medical Research Council (MRC)/ UK Department for International Development (DFID) under the MRC/DFID Concordat agreement.

## Data Availability

Data extracted from published studies is provided in the Supplementary Material (A1 Studies & A2 Studies). Unpublished data that support the findings of this study are available from the corresponding author upon reasonable request and subject to agreement with the data owners.

