## Supplementary Material for "Systematic review of the entomological impact of insecticide-treated nets evaluated using experimental hut trials in Africa"

### Supplementary methods: Systematic review process

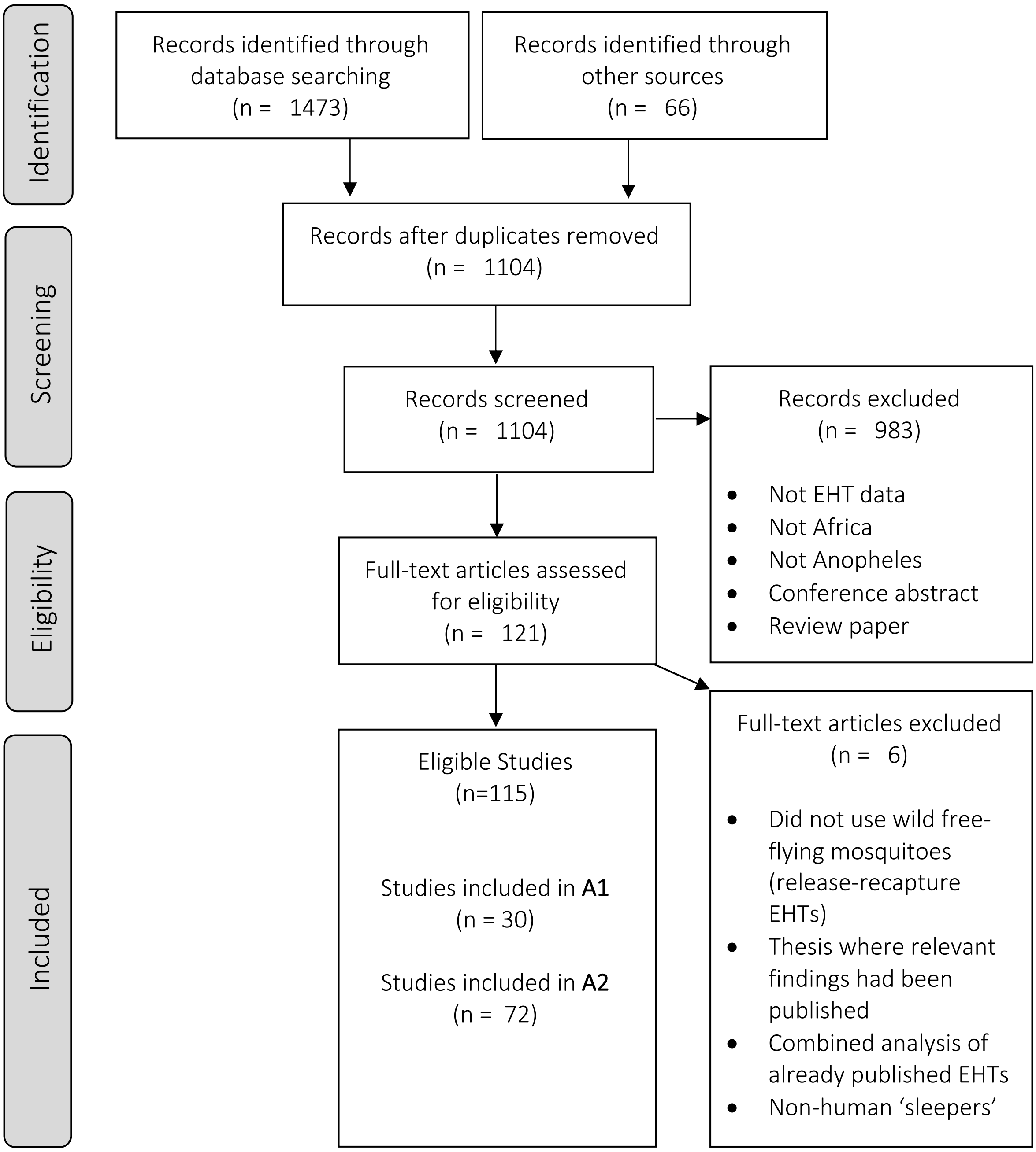


**Supplementary Figure 1.** PRISMA flow diagram to illustrate the phases of the systematic meta-analysis process. The protocol for the review was prospectively registered on PROSPERO (CRD42019117858).

The systematic search sought to collect data on all experimental hut trials (EHTs) investigating insecticide treated nets (ITNs) – encompassing conventionally treated nets (CTNs) and long-lasting insecticidal nets (LLINs) – and indoor residual spraying (IRS). The aim was to create a large database of all known EHTs to date investigating the two most common vector control interventions for malaria. As a systematic meta-analysis of IRS EHT data was recently completed by Sherrard-Smith et al[1], the main analysis of this paper focused on ITNs, with the intention for the database to continue to be updated with EHT data over time and form the basis for future analyses.

Supplementary Figure 1 shows the systematic process that was followed to identify and select the studies included in the database and which of these were then included in each meta-analysis. The search strings used were adapted for each database, generally following the outline for the MEDLINE database as described in Supplementary Table 1.

**Supplementary Table 1** – Search strings used in MEDLINE search.

| Search | Search term |
|---|---|
| 9 | 7 and 8 |
| 8 | malaria.mp. [mp=title, abstract, original title, name of substance word, subject heading word, floating sub-heading word, keyword heading word, protocol supplementary concept word, rare disease supplementary concept word, unique identifier, synonyms] |
| 7 | 5 and 6 |
| 6 | (hut or huts).mp. [mp=title, abstract, original title, name of substance word, subject heading word, floating sub-heading word, keyword heading word, protocol supplementary concept word, rare disease supplementary concept word, unique identifier, synonyms] |
| 5 | 1 or 2 or 3 |
| 3 | (insecticide treated net* or ITN* or long lasting insecticidal net* or LLIN*).mp. [mp=title, abstract, original title, name of substance word, subject heading word, floating sub-heading word, keyword heading word, protocol supplementary concept word, rare disease supplementary concept word, unique identifier, synonyms] |
| 2 | Insecticide-Treated Bednets/ |
| 1 | (indoor adj2 residual adj2 spray*) or IRS).mp. [mp=title, abstract, original title, name of substance word, subject heading word, floating sub-heading word, keyword heading word, protocol supplementary concept word, rare disease supplementary concept word, unique identifier, synonyms] |

The inclusion and exclusion criteria are shown in Supplementary Table 2. It is important to note that information was extracted for EHTs if data on mosquito outcomes (mortality, feeding, exiting) was not reported in the paper, as long as they fulfilled all other criteria. This is to ensure that a record was kept of all known trials in case contact could be made with the respective authors for the data. Overall, there are 115 EHT studies in the database, 30 of which were included in analysis 1 (A1) and 72 were included in analysis 2 (A2).

**Supplementary Table 2** – Inclusion and exclusion criteria used in the systematic review.

| Inclusion | Exclusion |
|---|---|
| Study conducted in Africa | Review paper |
| Must have control data | No control data |
| Must be investigating long-lasting insecticidal nets, indoor residual spraying or both of these interventions combined | No sleeper rotation |
| Studies must report mosquito mortality data* | Not a human sleeper |
| | Not Anopheles |
| | Not an EHT conducted using a WHO compliant protocol |

* *Information on studies was still extracted if data reported on mortality, blood feeding, exiting and total number of mosquitoes collected was incomplete. Studies were only included in analyses if mosquito mortality and other data relevant to the specific analysis was available. Contact was made with frequent EHT authors in order to try to obtain unpublished data.*

### Studies included in A1: Pyrethroid resistance vs EHT survival

**Supplementary Table 3.** List of studies where a concurrent pyrethroid discriminating dose bioassay was carried out. These studies were used to investigate the association between survival exhibited in pyrethroid resistance bioassays and experimental hut trials of ITNs. * = Studies where the Ifakara hut design was used. These studies were excluded from A1 (but data from these trials is included in Supplementary Figure 4).

| Study | No. of EHTs | Reference | Study site |
|---|---|---|---|
| 1 | 2 | Agossa et al. (2014) | Akron, Benin<br>Malanville, Benin |
| 2 | 1 | Asale et al. (2014) | Gilgel Gibe hydropower dam area, Ethiopia |
| 3 | 1 | Bayili et al. (2017) | Vallée du Kou, Burkina Faso |
| 4 | 3 | Corbel et al. (2010) | Malanville, Benin<br>Vallée du Kou, Burkina Faso<br>Pitoa, Cameroon |
| 5 | 1 | Ketoh et al. (2018) | Kolokopé, Togo |
| 6 | 1 | Koudou et al. (2011) | Yaokoffikro, Côte d'Ivoire |
| 7 | 1 | Kweka et al. (2017) | Lower Moshi, Tanzania |
| 8 | 1 | Malima et al. (2013) | Muheza, Tanzania |
| 9 | 1 | Malima et al. (2008) | Muheza, Tanzania |
| 10 | 2 | Mosha et al. (2008) | Lower Moshi, Tanzania |
| 11 | 1 | N'Guessan et al. (2016) | Cové, Benin |
| 12 | 1 | Ngufor et al. (2014a) | Tiassalé, Côte d'Ivoire |
| 13 | 1 | Ngufor et al. (2017) | Cové, Benin |
| 14 | 1 | Ngufor et al. (2016) | Cové, Benin |
| 15 | 2 | Ngufor et al. (2014b) | Vallée du Kou, Burkina Faso<br>Lupiro, Tanzania |
| 16* | 2 | Okumu et al. (2013) | Lupiro, Tanzania |
| 17 | 1 | Oxborough et al. (2013) | Lower Moshi, Tanzania |

| | | | |
|---|---|---|---|
| 18 | 1 | Pennetier et al. (2013) | Malanville, Benin |
| 19 | 2 | Toe et al. (2018) | Vallée du Kou, Burkina Faso<br>Tengrela, Burkina Faso |
| 20 | 1 | Tungu et al. (2010) | Muheza, Tanzania |
| 21* | 1 | Moore et al. (2016) | Ifakara, Tanzania |
| 22 | 1 | Ngufor et al. (2016) | Cové, Benin |
| 23 | 1 | Tungu et al. (2016) | Muheza, Tanzania |
| 24 | 1 | Rowland et al. (unp.) | Moshi, Tanzania |
| 25 | 1 | Rowland et al. (unp.) | Muheza, Tanzania |
| 26 | 1 | Guillet et al. (2008) | Yaokoffikro, Côte d'Ivoire |
| 27 | 1 | Miller et al. (1991) | Wali Kunda, The Gambia |
| 28 | 1 | Mosha et al. (2008) | Mabogini, Tanzania |
| 29 | 1 | Darriet et al. (1999) | Yaokoffikro, Côte d'Ivoire |
| 30 | 1 | Ngufor et al. (unp.) | Cové, Benin |

### Studies included in A2: Mosquito behaviour as survival increases

**Supplementary Table 4.** List of studies included in meta-analysis A2, which quantifying the impact of ITNs on different mosquito behaviours.
• = study included, ○ = study included but allocated a total number of mosquitoes (half the median) in order to include data that was provided as proportion fed.

| Study | No. of EHTs | Reference | Study site | Figure B | Figures C and D |
|---|---|---|---|---|---|
| 1 | 2 | Agossa et al. (2014) | Akron, Benin<br>Malanville, Benin | • | • |
| 2 | 1 | Badolo et al. (2012) | Pissy, Burkina Fasso | • | • |
| 3 | 1 | Bayili et al. (2015) | Vallée du Kou, Burkina Faso | • | • |
| 4 | 1 | Camara et al. (2018) | M'bé, Côte d'Ivoire | • | • |
| 5 | 3 | Corbel et al. (2010) | Malanville, Benin<br>Vallée du Kou, Burkina Faso<br>Pitoa, Cameroon | • | • |
| 6 | 1 | Djenontin et al. (2018) | Malanville, Benin | • | • |
| 7 | 1 | Ketoh et al. (2018) | Kolokopé, Togo | • | • |
| 8 | 1 | Koffi et al. (2015) | M'bé, Côte d'Ivoire | • | • |
| 9 | 1 | Kweka et al. (2017) | Lower Moshi, Tanzania | • | • |
| 10 | 1 | Mahande et al. (2018) | Magugu, Tanzania | • | • |
| 11 | 1 | Malima et al. (2013) | Muheza, Tanzania | • | • |
| 12 | 1 | N'Guessan et al. (2010) | Akron, Benin | • | • |
| 13 | 1 | N'Guessan et al. (2016) | Cové, Benin | • | • |
| 14 | 1 | Ngufor et al. (2016) | Cové, Benin | • | • |
| 15 | 1 | Pennetier et al. (2013) | Malanville, Benin | • | • |
| 16 | 1 | Tungu et al. (2010) | Muheza, Tanzania | • | • |
| 17 | 1 | Tungu et al. (2012) | Muheza, Tanzania | • | • |

| | | | | | |
|---|---|---|---|---|---|
| 18 | 1 | Rowland et al. (unp.) | Moshi, Tanzania | • | • |
| 19 | 1 | Rowland et al. (unp.) | Muheza, Tanzania | • | • |
| 20 | 1 | Adeogun et al. (2012) | New Bussa, Nigeria | • | • |
| 21 | 1 | Allossogbe et al. (2017) | Cové, Benin | • | • |
| 22 | 1 | Asale et al. (2014) | Gilgel Gibe hydropower dam area, Ethiopia | • | • |
| 23 | 1 | Chandre et al. (2010) | Vallée du Kou, Burkina Faso | • | • |
| 24 | 1 | Malima et al. (2008) | Muheza, Tanzania | • | • |
| 25 | 2 | Mosha et al. (2008) | Lower Moshi, Tanzania | • | • |
| 26 | 1 | N'Guessan et al. (2001) | Yaokoffikro, Côte d'Ivoire | • | • |
| 27 | 1 | Ngufor et al. (2014a) | Tiassalé, Côte d'Ivoire | • | • |
| 28 | 1 | Ngufor et al. (2017) | Cové, Benin | • | • |
| 29 | 1 | Ngufor et al. (2014c) | Akron, Benin | • | • |
| 30 | 2 | Ngufor et al. (2014b) | Vallée du Kou, Burkina Faso<br>Lupiro, Tanzania | • | • |
| 31 | 2 | Toe et al. (2018) | Vallée du Kou, Burkina Faso<br>Tengrela, Burkina Faso | • | • |
| 32 | 1 | Graham et al. (2005) | Muheza, Tanzania | | ○ |
| 33 | 1 | Koudou et al. (2011) | Yaokoffikro, Côte d'Ivoire | • | |
| 34 | 1 | N'Guessan et al. (2010) | Ladji, Benin | • | • |
| 35 | 1 | N'Guessan et al. (2009) | Ladji, Benin | • | • |
| 36 | 1 | N'Guessan et al. (2007) | Malanville, Benin | • | • |
| 37 | 1 | Oxborough et al. (2013) | Moshi, Tanzania | • | • |
| 38 | 1 | Winkler et al. (2012) | M'bé, Côte d'Ivoire | • | • |
| 39 | 1 | Tungu et al. (2017) | Muheza, Tanzania | | ○ |
| 40 | 1 | Bayili et al. (2017) | Vallée du Kou, Burkina Faso | | ○ |
| 41 | 1 | Tungu et al. (2017) | Muheza, Tanzania | | ○ |
| 42 | 1 | N'Guessan et al. (2016) | M'bé, Côte d'Ivoire | • | • |
| 43 | 1 | Ngufor et al. (2016) | Cové, Benin | • | • |
| 44 | 1 | Tungu et al. (2016) | Muheza, Tanzania | • | • |
| 45 | 1 | Koffi et al. (2015) | M'bé, Côte d'Ivoire | | ○ |
| 46 | 1 | Koffi et al. (2015) | M'bé, Côte d'Ivoire | | ○ |
| 47 | 1 | N'Guessan et al. (2015) | M'bé, Côte d'Ivoire | | ○ |
| 48 | 1 | Kweka et al. (2015) | Moshi, Tanzania | | ○ |
| 49 | 1 | Tungu et al. (2015) | Muheza, Tanzania | | ○ |
| 50 | 1 | Tungu et al. (2015) | Muheza, Tanzania | | ○ |
| 51 | 1 | N'Guessan et al. (2015) | M'bé, Côte d'Ivoire | | ○ |
| 52 | 1 | Tungu et al. (2013) | Muheza, Tanzania | | ○ |
| 53 | 1 | Sidick et al. (2011) | Malanville, Benin | | ○ |
| 54 | 1 | Chabi et al. (2009) | Malanville, Benin | | ○ |
| 55 | 1 | Tungu et al. (2009) | Muheza, Tanzania | | ○ |
| 56 | 1 | Dabire et al. (2008) | Vallée du Kou, Burkina Faso | | ○ |
| 57 | 1 | Tungu et al. (2008) | Muheza, Tanzania | | ○ |

| | | | | | |
|---|---|---|---|---|---|
| 58 | 1 | Chouaibou et al. (2006) | Pitoa, Cameroon | • | • |
| 59 | 1 | Corbel et al. (2004) | Contonou, Benin | • | • |
| 60 | 2 | Fane et al. (2012) | Koumantou, Mali<br>Sélingué, Mali | • | • |
| 61 | 1 | Fanello et al. (1999) | Yaokoffikro, Côte d'Ivoire | • | • |
| 62 | 1 | Guillet et al. (2008) | Yaokoffikro, Côte d'Ivoire | • | • |
| 63 | 2 | Hougard et al. (2003) | M'bé, Côte d'Ivoire<br>Yaokoffikro, Côte d'Ivoire | • | • |
| 64 | 1 | Kitau et al. (2014) | Moshi, Tanzania | • | • |
| 65 | 1 | Kolaczinski et al. (2000) | Yaokoffikro, Côte d'Ivoire | • | • |
| 66 | 1 | Malima et al. (2009) | Moshi, Tanzania | • | • |
| 67 | 1 | Miller et al. (1991) | Wali Kunda, The Gambia | • | • |
| 68 | 2 | Mosha et al. (2008) | Mabogini, Tanzania | • | • |
| 69 | 1 | Darriet et al. (1999) | Yaokoffikro, Côte d'Ivoire | • | • |
| 70 | 2 | Sanou et al. (unp.) | Tengrela, Burkina Faso<br>Tiefora, Burkina Faso | • | • |
| 71 | 1 | Tungu et al. (unp.) | Muheza, Tanzania | • | • |
| 72 | 1 | Ngufor et al. (unp.) | Cové, Benin | • | • |

### Supplementary information on the EHT database

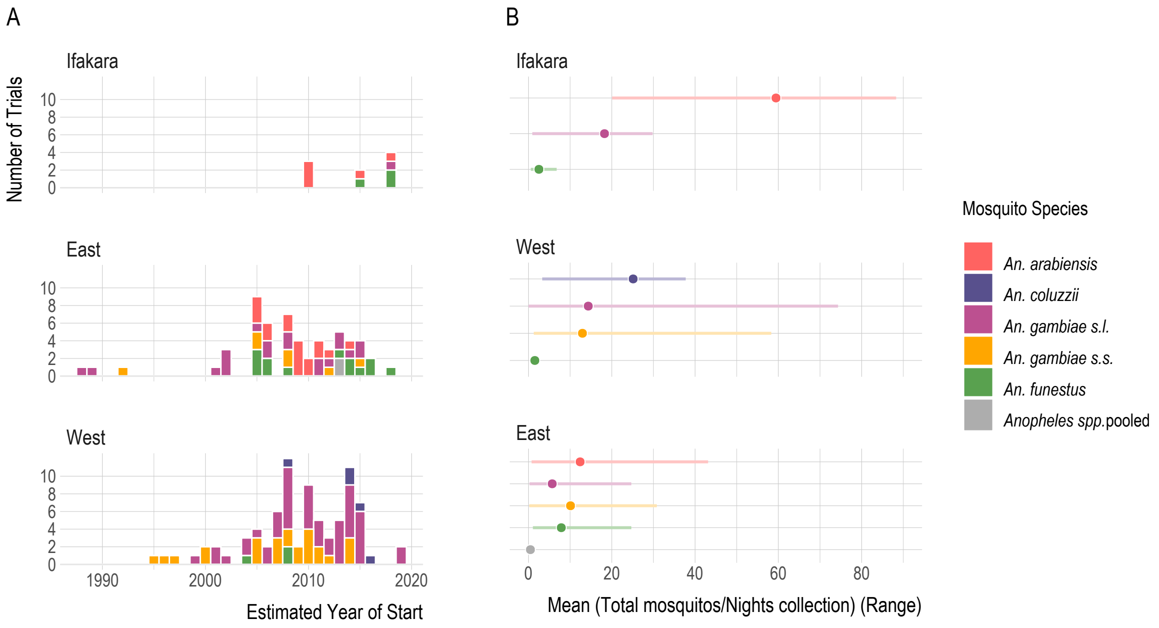


**Supplementary Figure 2.** (A) The number of experimental hut trials summarised by the estimated year of start, hut design and mosquito species. Trials are counted more than once if they presented data for multiple mosquito species. (B) Mean and range of the number of mosquitoes entering control huts of each design per night, as estimated from the total number of mosquitoes collected in the trial over the number of nights of collection.

The earliest trial identified used the East African hut design and took place in 1988, whereas the first trials identified using the West African and Ifakara designs were in 1995 and 2010 respectively (Supplementary Figure 2A). The greatest number of trials were carried out in 2008 (n=16), three of which presented data for multiple mosquito species.

Supplementary Figure 2B shows that there was substantial variation in the number of mosquitoes collected per night, particularly in Ifakara and West African huts. In east Africa, the mean number of *An. arabiensis* caught per night in East African and Ifakara hut designs was greater than for other species, but the range was also larger. The mean number of *An. funestus* collected per night was consistently low in all hut designs. Overall, Ifakara huts collected the most mosquitoes per night, followed by West African then East African huts.

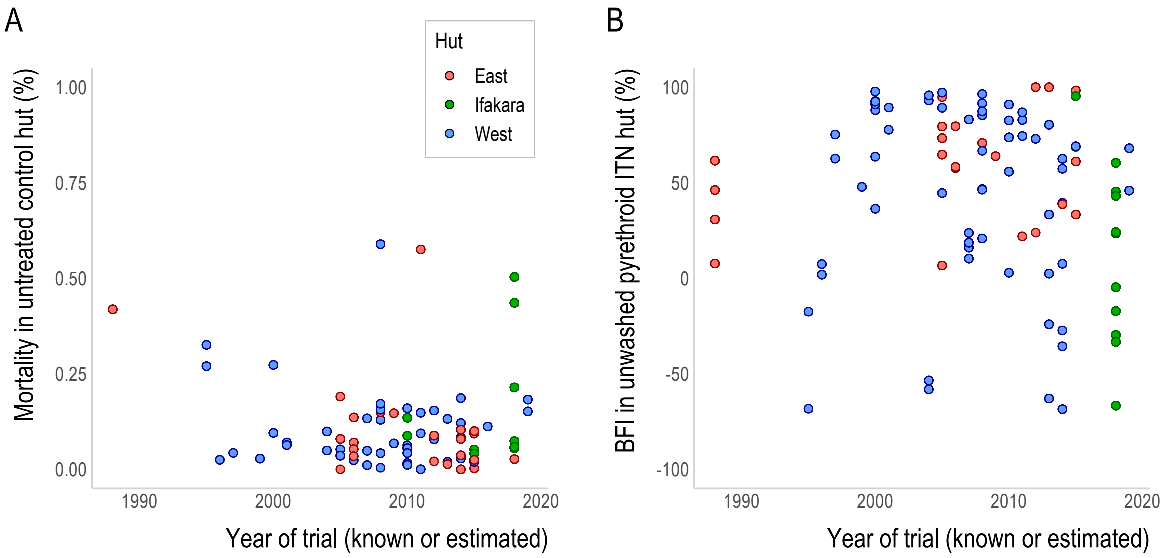


**Supplementary Figure 3.** (A) The percentage mosquito mortality exhibited in EHT arms evaluating untreated nets over time. (B) The percentage blood-feeding inhibition for mosquitoes in unwashed pyrethroid-only ITN huts over time. (Two datapoints that were below -100% were removed to aid visual inspection of these data.) If the trial start date was not stated, then it was estimated using the average time between known trial dates and their publication. Simple summary statistics for linear and logistic regression are provided in Supplementary Table 5. Plots are not included within figures as it is unlikely there has been a consistent change over time.

There was no evidence of a change in mortality in mosquitoes entering the control huts (Supplementary Figure 3A). An initial exploratory analysis of the raw data across trials revealed significant decreasing trends in the proportion of mosquitoes being inhibited from blood-feeding in pyrethroid-only ITN huts over time (Supplementary Figure 3B, Supplementary Table 5).

**Supplementary Table 5.** Summary statistics for (A) the logistic regression of trial date and mortality in pyrethroid-only ITN huts and (B) the linear regression of trial date and blood-feeding inhibition in pyrethroid-only ITN huts. Time was included as the dependent variable in all plots with no other covariates included. Significance of the time variable (p-value) was estimated using a likelihood ratio test.

| **Mortality in pyrethroid-only ITN huts over time** | | | | |
|---|---|---|---|---|
| | Estimate | Std. Error | Z value | p- value |
| Intercept | 34.67 | 1.40 | 24.78 | <2e-16 |
| Trial Year | -0.02 | <0.01 | -25.69 | <2e-16 |
| **Blood-feeding inhibition in pyrethroid-only ITN huts over time** | | | | |
| | Estimate | Std. Error | Z value | p- value |
| Intercept | 60.86 | 7.03 | 8.66 | <2e-16 |
| Trial Year | -0.03 | <0.01 | -8.64 | <2e-16 |

### Supplementary information on A1: Pyrethroid resistance vs EHT survival

Analyses A1 used data from EHTs investigating pyrethroid-only ITNs, where mosquito mortality in EHTs and concurrent pyrethroid susceptibility bioassays were carried out. Mosquitoes collected in the 3 eligible trials using the Ifakara hut design exhibited consistently low levels of EHT mortality, regardless of the level of insecticide resistance measured in the wild mosquito population (Supplementary Figure 4). Due to the outlying trend shown by the data, the small number of eligible trials, and Ifakara huts only being used in one trial site, it was decided to exclude Ifakara trials from further analysis.

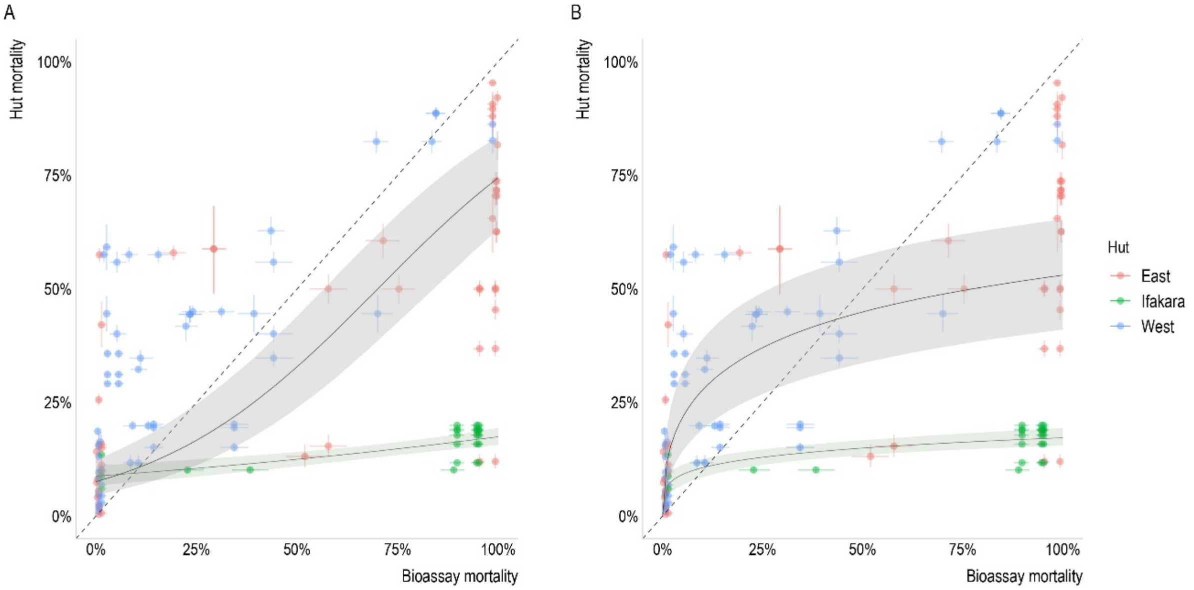


**Supplementary Figure 4. Comparison of models fit to East and West African huts designs (combined) and Ifakara.** A) Logistic model and B) Log-logistic model. Plots to illustrate the disparity concerning the predicted relationship between mortality observed in pyrethroid resistance bioassays and experimental hut trials in East and West African hut designs (grey) and the Ifakara hut design (green).

### Supplementary information on A2: Association between different EHT outcomes

#### Alternative function considered for deterrence

In addition to a three-parameter curved function for deterrence, a simple linear function with only one parameter was also considered (Supplementary Table 6). The expected log pointwise predictive density (elpd) values - obtained from leave-one-out cross validation - evaluates the goodness of fit of the statistical model to the data. These values suggest that the fit for option 1 is very slightly favoured, but the fits are very similar.

**Supplementary Table 6. Comparison of different functions for deterrence.** The two options considered for the functional relationship between deterrence and hut trial survival ($\theta_H^i$). The expected log pointwise predictive density (elpd) values describe the goodness of fit to all data irrespective of hut design and the fits to East African and West African trial data separately (with higher values indicating better goodness of fit). Option 1 is a three-parameter ($\delta_1, \delta_2, \delta_3$) function, whereas option 2 is a single parameter ($\delta_1$) function.

| Equation describing the change in deterrence with increasing hut survival | Elpd value (leave-one-out cross-validation) | | |
|---|---|---|---|
| | *All data* | *East only* | *West only* |
| Option 1 – Used in A2<br>$g(\theta_H^i, \delta_1, \delta_2, \delta_3) = \delta_1(\exp(\delta_2\,(1 - \exp(\delta_3 \theta_H^i))/\delta_3))$ | *-4314.2* | *-1185.5* | *-3110.7* |
| Option 2 – Not used in A2<br>$g(\theta_H^i, \delta_1) = \delta_1\,(1 - \theta_H^i)$ | *-4314.6* | *-1117.0* | *-3197.2* |

The ratio between the number of mosquitoes collected in control huts and intervention huts fit using option 1 and option 2 are displayed in Supplementary Figure 5A and 5D respectively. They both show very similar fits, but for option 2, the ratio of the number of mosquitoes collected in control huts compared to intervention huts falls below zero beyond 85% hut survival - suggesting that greater numbers of mosquitoes are collected from intervention huts compared to control huts. The probability of deterrence for each option is shown in Figures 5B and 5E. For option 1, although deterrence declines it doesn't fall to zero when the probability on hut survival is high (Figure 5B). Option 2 fixes the probability of hut survival to 1 when the probability of deterrence is zero, therefore the 95% credible interval for deterrence is very small when the probability of hut survival is high (Figure 5E). Ultimately, to allow the model more flexibility and more realistic uncertainty when the probability of hut survival is high, option 2 was not chosen.

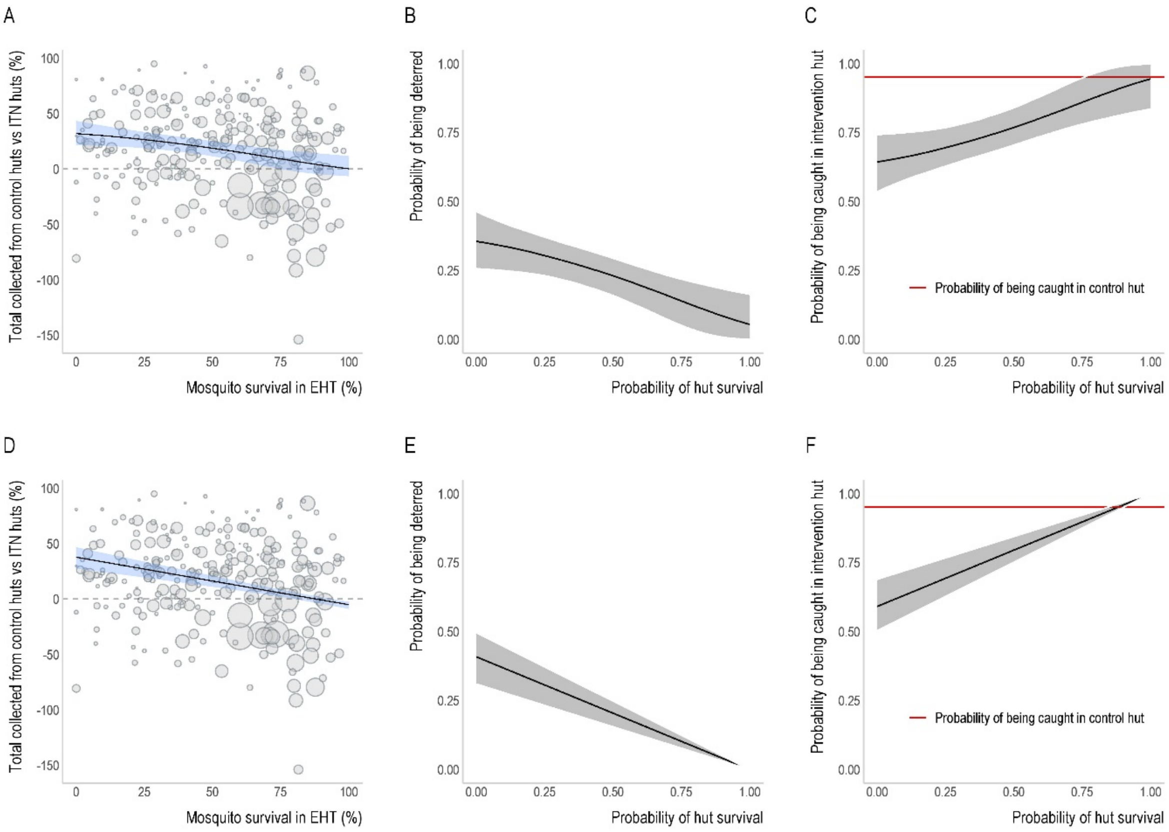


**Supplementary Figure 5. The two options considered for the functional relationship between deterrence and hut trial survival described in Supplementary Table 6.** A) Option 1 and D) Option 2 fit to all data irrespective of hut design. B-C and E-F) The probability of being deterred and the probability of being caught in an intervention hut as the probability of hut survival increases for Option 1 and Option 2 respectively. The red line in panels C and F depicts the constant probability of being caught in a control hut, irrespective of hut design and the level of hut survival.

### The probability of deterrence for East and West African hut designs

Supplementary Figure 6 shows the separate estimates for the probability of deterrence and the ratio between the number of mosquitoes collected in control huts and intervention huts for data from East and West African hut designs.

When the model is fit to East African data, although deterrence declines, there is very little change in the probability of deterrence as the probability of hut survival increases. However, there are fewer data points for the number of mosquitoes collected at greater levels of hut survival for East African huts, which may influence the fit. The West African fit is more similar to the combined fit to all data, simply reflecting that West African trials make up the bulk of the data. However, at ~80% hut trial survival the ratio falls below zero, indicating that more mosquitoes are collected from intervention huts than control huts at high survival. There is considerable variability in the number of mosquitoes caught in the control versus intervention huts and how this changes with hut survival. This difference in deterrence between studies with the same hut design appears greater than the differences between designs.

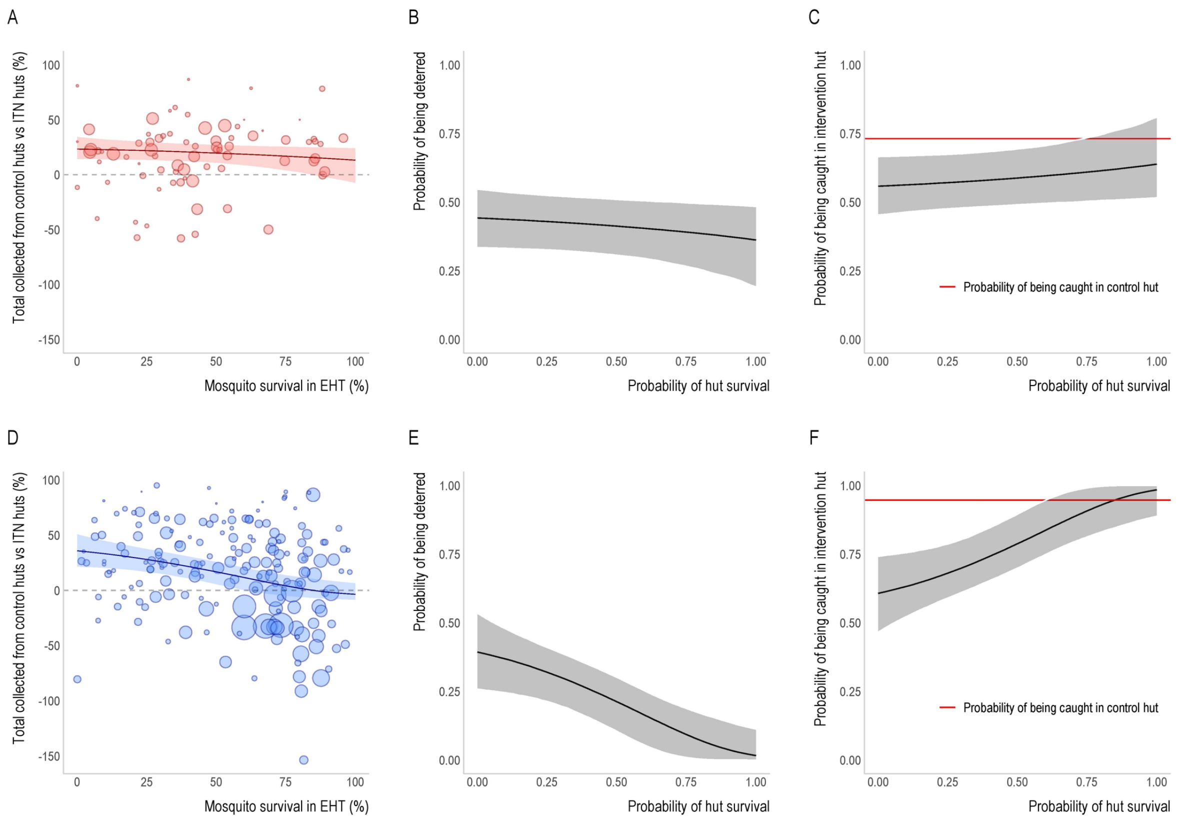


**Supplementary Figure 6. Difference in deterrence models for the two hut designs.** The ratio between the number of mosquitoes collected from control huts compared to intervention huts for A) East African trials and D) West African trials. B-C and E-F) The probability of being deterred and the probability of being caught in an intervention hut as the probability of hut survival increases for East African and West African huts respectively. The red lines in panels C and F depict the constant probability of being caught in a control hut of each design, irrespective of the level of hut survival.

### The probabilities of each outcome of a mosquito feeding attempt using the logistic fit

To predict the impact of resistance on the probability of each outcome of a mosquito feeding attempt, the logistic and log-logistic models were fit to data from each hut design separately. The log-logistic fit by hut design is shown in the main paper (Figure 4) and the logistic fit is shown here in Supplementary Figure 7. Panel A shows the logistic relationship between hut survival and bioassay survival by hut design. Lines are broadly similar with mosquitoes in West African huts generally having a higher level of survival for the same level of pyrethroid resistance. Similar to the predictions using the log-logistic fit, the probability of a mosquito successfully blood-feeding in an East African hut remains very low as resistance increases, only reaching 3.5% (Supplementary Figure 7B). Whereas, the probability in West African huts rises from 1% to 52% in a highly resistant population (Supplementary Figure 7C). The probability of being deterred appears high in mosquitoes attempting to feed in East huts at all levels of resistance, only reducing by 5% to 38%. Deterrence falls much lower for West huts, declining from 31% in a susceptible population to 3% in a highly resistant population. The probability of exiting unfed from West huts rises from 17% to 45% before declining to 39% at high resistance. Whilst the probability of exiting unfed from East huts increases 38% to 52%.

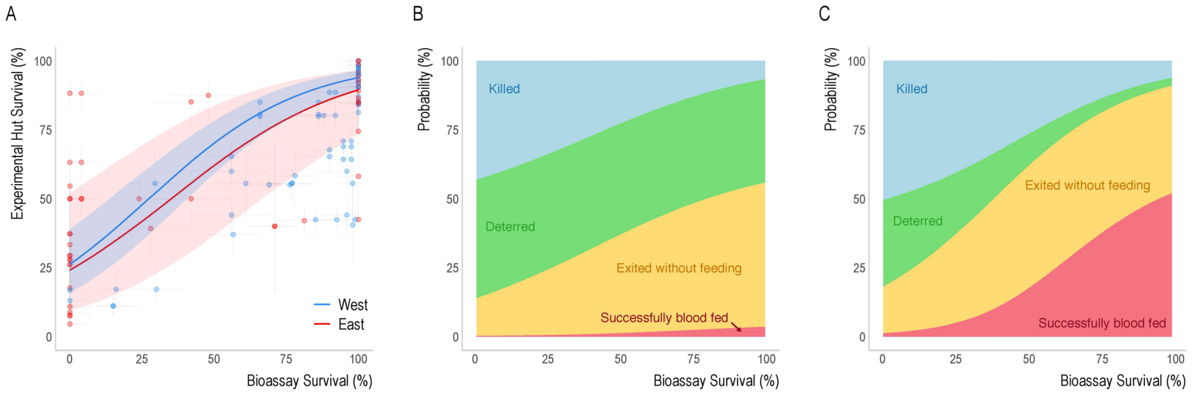


**Supplementary Figure 7. Summary of the difference between East and West African hut design and how this is predicted to change the outcome of a single blood-feeding event.** A) The logistic relationship between experimental hut trial survival and bioassay survival for trials of each hut design. B-C) Average probability that blood feeding mosquitoes will be killed, exit without feeding, be deterred from entering or successfully blood feed in B) East African huts or C) West African huts, assuming that the relationship between hut trial mortality and bioassay mortality is determined by the logistic model (see Figure 4 of the main text for other model fits).

### Model comparisons

Models were fit to data for all ITNs identified in the systematic review irrespective of the insecticide used. This make the assumption that though the mortality induced by the ITN might vary, the relationship between mortality and other factors influencing ITN efficacy (i.e. blood-feeding and deterrence) remains constant. These models were compared to models fit to restricted data for pyrethroid-only, pyrethroid-only and pyrethroid-PBO or pyrethroid-only and pyrethroid-combination nets (Supplementary Table 7). Models were evaluated by comparing the fit of the different models (using elpd) to a common dataset (i.e. the pyrethroid-only ITN data). The difference in elpd between the models fit with the reduced and full dataset is sufficiently small (relative to the magnitude of the elpd) to justify the use of the full dataset which are presented in the main text.

**Supplementary Table 7. Table comparing the model fits for models fit with different datasets.** Models were fit using four datasets: 1) All nets: Including data for all ITNs irrespective of the insecticide on the net (all other datasets are nested within these data), 2) Pyrethroid-only nets: All ITNs incorporating pyrethroid insecticide only, 3) Pyrethroid-only and pyrethroid-combination nets: All pyrethroid-only ITNs and pyrethroid combined with an alternative insecticide or the synergist piperonyl butoxide (PBO) and 4) Pyrethroid-only ITNs and pyrethroid-PBO ITNs. Model comparisons were performed by estimating the goodness of fit to data common between all four models (pyrethroid-only ITN data).

| **Deterrence models** | | | | | |
|---|---|---|---|---|---|
| Model | Elpd_diff | Se_diff | Elpd_loo | P_loo | looic |
| 3 | 0.0 | 0.0 | -2436.2 | 238.7 | 4872.4 |
| 1 | -0.3 | 3.8 | -2436.5 | 241.2 | 4873.0 |
| 4 | -3.9 | 3.8 | -2440.1 | 238.9 | 4880.2 |
| 2 | -8.3 | 5.0 | -2444.5 | 232.0 | 4889.0 |
| **Successful blood-feeding models** | | | | | |
| Model | Elpd_diff | Se_diff | Elpd_loo | P_loo | looic |
| 2 | 0.0 | 0.0 | -1914.7 | 299.7 | 3829.4 |
| 4 | -4.3 | 18.8 | -1919.0 | 286.8 | 3838.1 |
| 3 | -17.8 | 38.9 | -1932.5 | 287.7 | 3865.1 |
| 1 | -38.0 | 47.9 | -1952.7 | 267.2 | 3905.4 |

**Supplementary Table 8. Summary of all functional relationships and best fit parameter values.** Table shows the functional relationships used in A1-2 and the mean parameter values from fitting all ITN data (either grouping east and west hut design together or separating dataset). Notation in this manuscript is compared to that previously used.

| **Probability** | **Notation as in Griffin 2010**[2] | **Mean parameter values** | | |
|---|---|---|---|---|
| | | All data | West huts only | East huts only |
| **Relationship between EHT survival ($\theta_H^i$) and susceptibility bioassay survival ($\theta_B^i$)**<br><br>Logistic fit:<br><br>$\theta_H^i = 1 - (1/(1 + \exp(-((1 - \theta_B^i) - \alpha)\beta)))$<br><br>Log-logistic fit:<br><br>$\theta_H^i = 1 - (1/(1 + ((1 - \theta_B^i)/\beta)^{-\alpha}))$ | $1 - l_p$ | $\alpha$ = 0.7<br>$\beta$ = 3.57<br><br>$\alpha$ = 0.47<br>$\beta$ = 0.89 | $\alpha$ = 0.73<br>$\beta$ = 3.78<br><br>$\alpha$ = 0.88<br>$\beta$ = 0.72 | $\alpha$ = 0.66<br>$\beta$= 3.29<br><br>$\alpha$ = 0.36<br>$\beta$ = 1.05 |
| **Relationship between EHT survival ($\theta_H^i$) and the probability of mosquitoes being deterred ($\theta_D^i$) from entering a hut with an ITN**<br><br>$\theta_D^i = \delta_1 (\exp(\delta_2 (1 - \exp(\delta_3 \theta_H^i))/\delta_3))$<br><br>Difference in the proportion of mosquitoes collected in control huts ($\theta_C^i$) relative to ITN huts ($\theta_I^i$) as survival increases:<br><br>$\theta_I^i = 1 - \theta_D^i$<br><br>$\delta_C = (\theta_C^i - \theta_I^i)/\theta_C^i$<br><br>*The probability of mosquitoes entering and being caught in control huts is assumed to remain constant, independent of the level of survival in the hut. | $m_1$ | $\delta_1$ = 0.36<br>$\delta_2$ = 0.49<br>$\delta_3$ = 2.57<br><br>$\theta_C^i$ = 0.95* | $\delta_1$ = 0.39<br>$\delta_2$ = 0.66<br>$\delta_3$ = 3.13<br><br>$\theta_C^i$ = 0.95 | $\delta_1$ = 0.45<br>$\delta_2$ = 0.07<br>$\delta_3$ = 1.76<br><br>$\theta_C^i$ = 0.73 |
| **Relationship between EHT survival ($\theta_H^i$) and the probability of successfully blood-feeding ($\theta_{SF}^i$) after entering a hut with an ITN**<br><br>$\theta_{SF}^i = 1 - (\exp(\delta_4(1 - \exp(\delta_5 \theta_H^i))/\delta_5))$ | $k_1$ | $\delta_4$= 0.04<br>$\delta_5$ = 4.66 | $\delta_4$= 0.04<br>$\delta_5$ = 5.13 | $\delta_4$= 0.01<br>$\delta_5$= 3.43 |

| | | |
|---|---|---|
| **Probability of exiting without feeding after entering a hut with an ITN ($\theta_{EU}^{i}$)** <br> $\theta_{EU}^{i} = 1 - \theta_{SF}^{i} - (1 - \theta_{H}^{i})$ | $j_1 = 1 - k_1 - l_1$ | Parameters defined above |
